# Bifurcation analysis of a two-infection transmission model with explicit vector dynamics

**DOI:** 10.1101/2023.12.28.23300607

**Authors:** Akhil Kumar Srivastav, Vanessa Steindorf, Bruno V. Guerrero, Nico Stollenwerk, Bob W. Kooi, Maíra Aguiar

## Abstract

The investigation of epidemiological scenarios characterized by chaotic dynamics is crucial for understanding disease spread and improving disease control strategies. Motivated by dengue fever epidemiology, in this study we introduce the SIRSIR-UV model, which accounts for differences between primary and secondary infections and explicit disease vector dynamics. Our analysis, employing nonlinear dynamics and bifurcation theory, provides key insights into how vectors contribute to the overall system dynamics. In this paper, the formalization of backward bifurcation using center manifold theory, computation of Hopf and global homoclinic bifurcation curves, and derivation of analytical expressions for transcritical and tangent bifurcations deepen the understanding. The observation of chaotic behavior with the inclusion of seasonal forcing in the vector population underscores the importance of considering external factors like climate in disease spread. Our findings align with those from previous models, emphasizing the significance of simplifying assumptions, such as implicit vector dynamics, when constructing models without vector control. This study brings significant insights to the mathematical modeling of vector-borne diseases, providing a manageable framework for exploring complex epidemiological scenarios and identifying key factors influencing disease spread. While the absence of strain structure may limit predictive power in certain scenarios, the SIRSIR-UV model serves as a starting point for understanding vector-borne infectious disease dynamics.

## 1. Introduction

Mathematical models describing dengue epidemiological dynamics go back to 1970 [16]. To date, various aspects of the disease have been studied through mathematical models [2, 1]. Different extensions of the classical SIR models including the well known immunological Antibody-Dependent Enhancement (ADE) factor has demonstrated critical fluctuations with power law distributions of disease cases [23] and deterministic chaos [5, 10, 20].

Several mathematical models describing the transmission of dengue viruses have been proposed to explain the irregular behavior of disease epidemics [15, 29, 38, 24, 27, 3]. Analysis of a minimalistic two-infection dengue model, with at least two different dengue serotypes to describe differences between primary and secondary infections, has shown deterministic chaos in wider and biological parameter regions not anticipated by previous models [5, 3, 6]. Consideration of temporary cross-immunity (TCI) together with ADE gave rise to different bifurcation structures up to chaotic attractors able to describe large fluctuations observed in empirical outbreak data [3, 6, 7]. Finding wide ranges of a complex dynamical behaviour in the dengue model has opened new ways to analyze the existing empirical data, indicating that complex behaviour (deterministic chaos) is much more important in public health epidemiological systems than previously thought. This finding enhances the understanding of the seasonal, though non-periodic pattern [3, 10, 6, 7, 19, 6], providing improved forecasting capabilities, even though for shorter periods [35]. Such studies have provided essential guidance for public health organizations [4, 9, 17].

From an analytical and theoretical perspective, the motivation of dengue fever epidemiology has also supported insights into the mechanisms required to generate complex dynamics in simple epidemiological models [34, 8, 32, 33, 31, 18]. However, the effect of the vector dynamics was not explicitly modeled but implicitly by seasonal infectivity.

In the analysis of a simplified version of the multi-strain dengue model proposed by Aguiar and colleagues [5], as discussed in [34], the authors examined a compartmental model structured as a two-infection SIRSIR model without strain specificity of the pathogens. The model includes temporary immunity gained after a primary infection and incorporates an enhancement factor for a secondary infection based on the Antibody-dependent Enhancement (ADE) process.

Continuing the analysis, where complex dynamics were identified [34], including transcritical (sub-critical and supercritical) bifurcations (commonly known as forward and backward bifurcations in epidemiology), Hopf and global homoclinic bifurcation curves, and the points where these curves intersect (Cusp and Bogdanov-Takens bifurcations), a step further is to introduce the explicit interaction with mosquito population into the model. Since dengue fever is transmitted to humans through the bite of an infected mosquito, this addition allows for a more comprehensive and biological representation of the host (human)-vector (mosquito)-pathogen cycle. Therefore, this work aims to couple the vector population dynamics in a straightforward manner, as described in [2], to the model proposed in [34, 8]. The primary goal is to analyze the role of vector dynamics in the changes of mathematical bifurcation structures found in the SIRSIR-type model describing two subsequent infections and no strain structure of pathogens. This analysis will provide insights into the importance of vector dynamics in modeling infectious diseases transmitted by host-vector contacts.

This paper unfolds as follows. Upon describing the deterministic dynamic model in Section 2, a detailed mathematical analysis is performed in Section 3, featuring the steady-state solutions and characterization of the main bifurcation structures found analytically. In Section 4, numerical simulations are used to illustrate the bifurcation structures, along with an extensive stability analysis of the endemic equilibria. The inclusion of seasonality, resembling the changes in mosquito abundance in the model, is studied in Section 5, revealing the appearance of chaotic structures. The concluding section offers insights into the significance of employing simplifying assumptions during the development of mathematical models.

## 2. The two-infection model with coupled vector dynamics

In this study we extend the model described in [34, 8] by incorporating the dynamics of a disease vector population. This extension explicitly involves the mosquito population, where, using standard incidence (as opposed to massaction incidence), susceptible human individuals become infected at a rate *β* upon encountering an infected mosquito. Conversely, susceptible mosquitoes become infected by biting an infected human at a rate *ϑ*.

The flowchart illustrating the two-infection model with coupled vector dynamics is presented in Figure 1, outlining the compartmental system of ordinary differential equations that describes the assumptions underlying dengue fever epidemiology.

**Figure 1.**
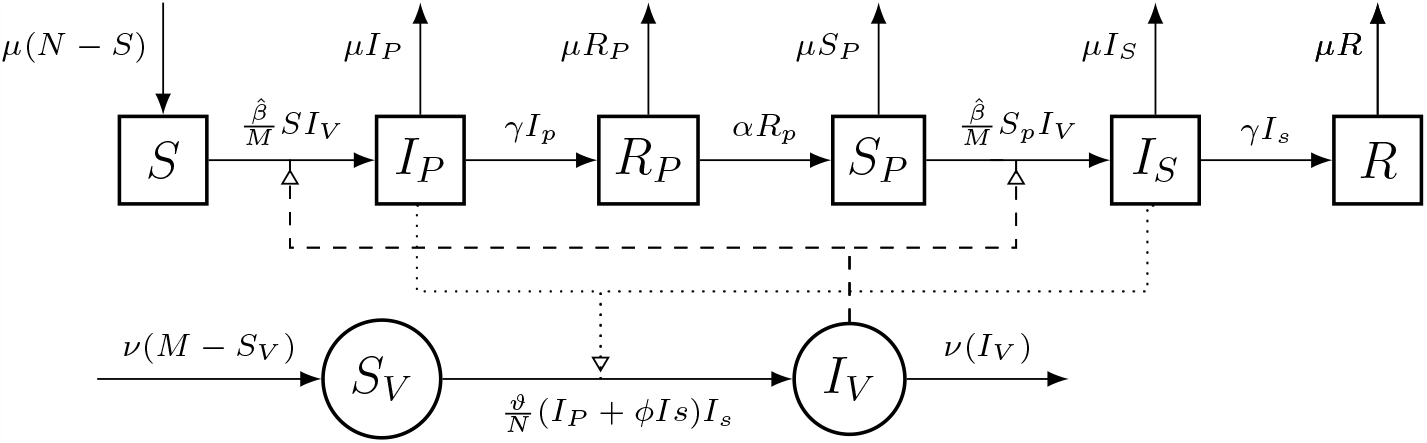
Flowchart for the compartmental model with two subsequent infections, temporary immunity, and vector dynamics. The arrows illustrate the flow of individuals among compartments based on their disease state. The square-shaped compartments are used to indicate states belonging to the host, while the circular ones are for those of the vector. The dotted and dashed lines represent the occurrence of disease transmission, indicating the flow from infected humans to susceptible vectors and from vectors to susceptible humans, respectively.

The set of ordinary differential equations (ODEs) for the deterministic two-infection epidemiological system, as an extension of [34] to include explicit vector dynamics, is given by:

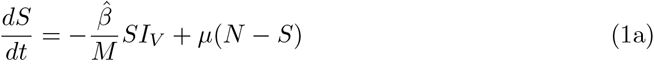

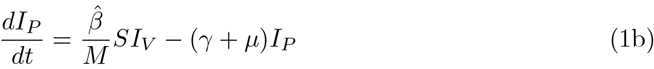

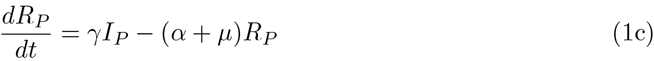

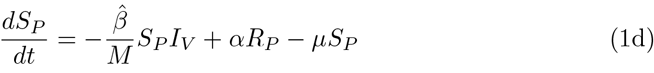

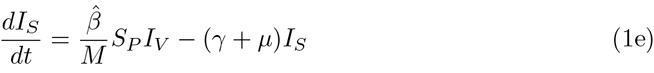

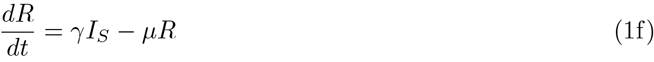

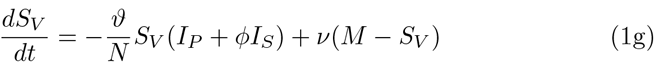

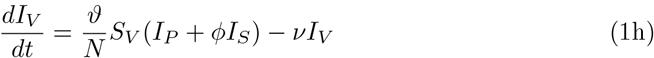

and the dynamics is described as follows.

Susceptible individuals (*S*) become infected for the first time (*I*_*P*_) through the bite of an infected mosquito (*I*_*V*_) at a rate 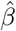. They then recover from the first infection (*R*_*P*_) after a period of 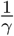 and gain temporary immunity to the second infection. After the temporary immunity period of 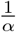, they become susceptible again (*S*_*P*_). Individuals who have experienced a previous dengue infection can be infected for the second time (*I*_*S*_) through the bite of an infected mosquito (*I*_*V*_) at the same rate 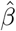. Finally, individuals develop long-term immunity after a recovery time of 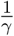.

On the other hand, susceptible mosquitoes (*S*_*V*_) can acquire the infection from individuals in either a primary or secondary infection. Hence, the force of infection is given by 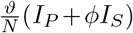. The constant ratio *ϕ* is introduced to account for the difference in transmissibility of the disease to vectors from individuals in primary and secondary infections. This factor is essential in light of the assumption that severe disease is associated with secondary dengue infection due to the antibody-dependent enhancement (ADE) process, as detailed in the prior study [34]. Consequently, *ϕ* acts as a scaling factor, allowing for the differentiation of the baseline infectivity rate (in this case, *ϑ*). The assumption, consistent with [34], is that if *ϕ >* 1, individuals in secondary infections transmit more due to a higher viral load. Conversely, if *ϕ <* 1, it means that individuals experiencing a secondary severe infection contribute less to the spread of the infection due to hospitalization, leading to reduced mobility and interactions.

Additionally, we make the assumption that natural mortality and birth rates are equal for individuals in both the host and vector populations, denoted as *μ* and *ν* respectively, irrespective of their disease state. Consequently, the dynamics for the total population, given by 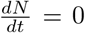, and for the vector population, given by 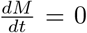, are satisfied. This implies that the total population, represented by *N* = *S* +*S*_*P*_ +*R*_*P*_ +*I*_*P*_ +*I*_*S*_ +*R*, and the vector population, represented by *M* = *S*_*V*_ + *I*_*V*_, remain constant over time.

In the subsequent sections, we analyze the host model with two infections and vector dynamics described in equation (1).

## 3. Analysis of the SIRSIR-UV type model

### 3.1. Positivity and boundedness of the solutions

In this section, we establish the positivity and boundedness of the solutions for the system (1). Examining the system (1), we observe the following properties:

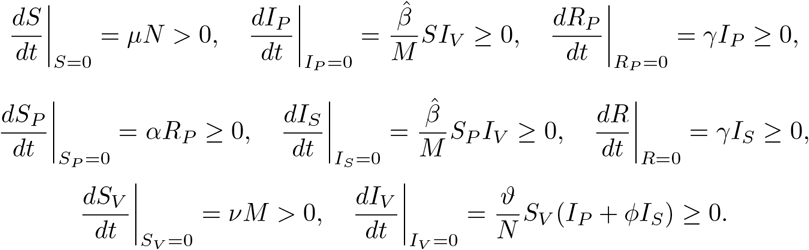

All parameters are non-negative constants, ensuring that all the mentioned inequalities are non-negative on the boundary planes (refer to [34] for further details). Consequently, the solutions of the system (1) remain non-negative. Furthermore, the total populations *N* and *M* are constant, as described in the previous section, because 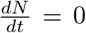 and 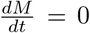. Therefore, the solutions *S, I*_*P*_, *R*_*P*_, *S*_*P*_, *I*_*S*_, *R* are bounded by *N*, and similarly, the solutions *S*_*V*_, *I*_*V*_ are bounded by *M*. Hence, the biologically feasible region of the model system (1) is defined by the following positive invariant set.

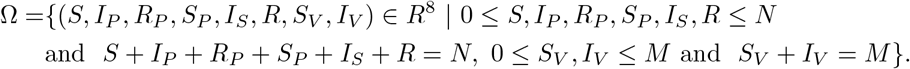

### 3.2. Steady state solutions

#### 3.2.1. Disease-Free Equilibrium (DFE) *E*_0_

The system (1) always possesses a disease-free equilibrium (DFE) within the region Ω, defined as

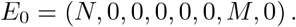

Concerning the DFE, it is customary to calculate the basic reproduction number (ℛ_0_). Mathematically, ℛ_0_ is computed using the next generation method, as described in [37]. This method has been applied to vector-borne disease models in [37, 30, 36] (refer to Appendix 6). The reproduction number serves as a threshold for the existence of an endemic equilibrium (positive steady state) and the stability of the DFE. Specifically, it indicates when the transcritical supercritical bifurcation occurs (see [34]).

Applying the next generation matrix method from [37], the basic reproduction number for the system (1) is given by

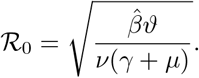

The calculation of the ℛ_0_. The detailed calculation of ℛ_0_ can be found in Appendix 6.

The epidemiological interpretation of ℛ_0_ is as follows. The term 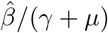 signifies the expected number of infected humans generated by a single infected human in a disease-free human population during their infectious period. Similarly, the term *ϑ/ν* denotes the expected number of infected mosquitoes generated by a single infected mosquito in a disease-free mosquito population during their infectious period. The square root of the product of these two terms, ℛ_0_, represents the number of secondary infected hosts generated by a single infected human and a single infected mosquito throughout the entire infectious period in a susceptible population.

Figure 2 illustrates the values of ℛ_0_ as a function of the parameters 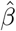, *ϑ*, and *γ*, representing the transmission rates for humans and vectors and the recovery rate, respectively. It can be observed that ℛ_0_ is a monotonically increasing function of 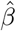and *ϑ*, while its value is inversely proportional to the parameter *γ*.

**Figure 2.**
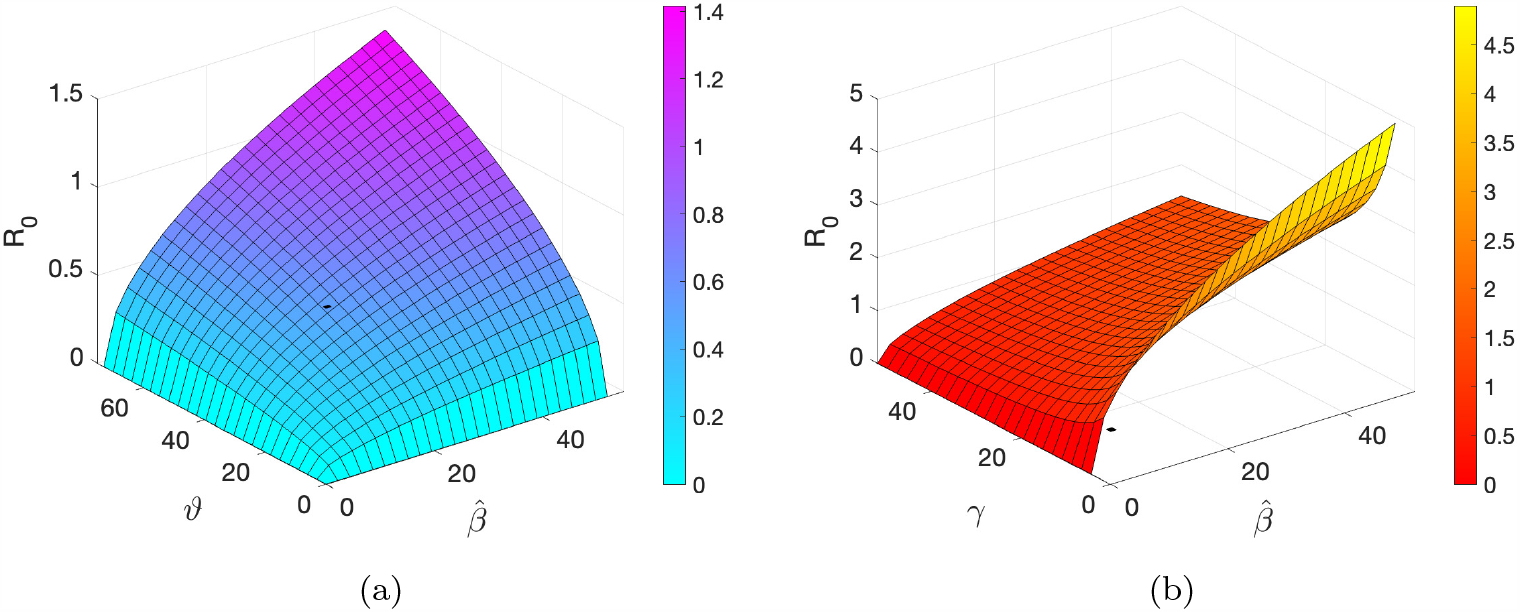
ℛ_0_ as function of the parameters 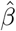, *ϑ, γ*.

#### 3.2.2. Disease-Endemic Equilibrium (DEE) *E*_1_

System (1) has an endemic equilibrium denoted by 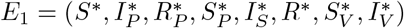. The endemic equilibrium satisfies the following algebraic qualities:

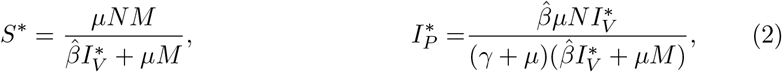

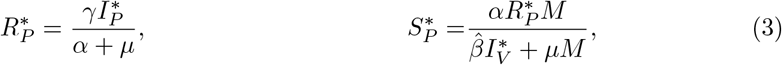

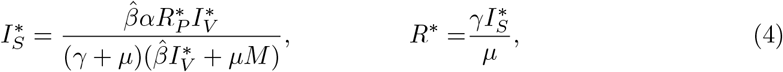

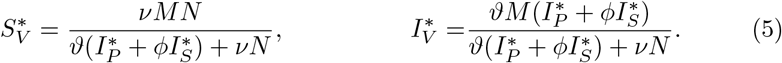

Note that all entries of the equilibrium 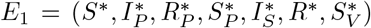 are described in terms of the model parameters and the variable *I*_*V*_. Furthermore, 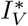 is the root of the following quadratic polynomial *P* :

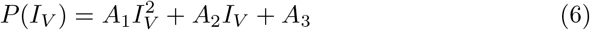

where the coefficients are explicitly written below in terms of the model parameters.

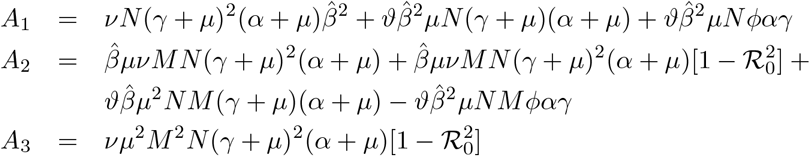

Therefore, *E*_1_ represents an equilibrium of system (1) if the roots of the polynomial *P* are positive. To analyze the positive roots, we apply Descartes’ rule of signs to equation (6), and the possible positive roots of the polynomial *P* are listed in Table 1.

**Table 1:** Descartes’ rules showing the number of positive roots for the polynomial *P*.

| Case $\mathcal{R}_0 > 1$ | $A_1$ | $A_2$ | $A_3$ | Changes of signs | Positive roots |
| --- | --- | --- | --- | --- | --- |
| i) | + | + | - | 1 | One |
| ii) | + | - | - | 1 | One |
| Case $\mathcal{R}_0 < 1$ | $A_1$ | $A_2$ | $A_3$ | Changes of signs | Positive roots |
| i) | + | + | + | 0 | None |
| ii) | + | - | + | 2 | Two |

Table 1 displays the potential number of positive roots for the polynomial. It is important to note that if ℛ_0_ *>* 1, only one positive root is possible. However, if ℛ_0_ *<* 1, there are two possibilities: two positive roots or none.

Therefore, we can conclude the following results.

##### Theorem 3.1.

*If ℛ*_0_ *>* 1, *then the system (1) has only one endemic equilibrium in the* Ω *region*.

*Proof*. Note that the coefficient of the polynomial *P, A*_1_, is always positive. Since ℛ_0_ *>* 1, then *A*_3_ *<* 0 (always negative). Thus, the polynomial has only one positive root, namely,

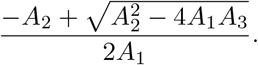

Therefore, a unique endemic equilibrium exists. □

##### Theorem 3.2.

*If ℛ*_0_ *<* 1, *then the system (1) has two positive endemic equilibria in the* Ω *region*.

*Proof*. Regardless of the specific values of the positive parameters, *A*_1_ *>* 0 always. If ℛ_0_ *<* 1, then *A*_3_ *>* 0 (always positive). Thus, the polynomial has either none or two positive roots.

In the case of two positive roots, it means that the system exhibits a backward bifurcation. That is, there are two endemic equilibria when ℛ_0_ *<* 1. Therefore, a sub-critical transcritical bifurcation can occur. □

To characterize this so-called backward bifurcation, in the next section, we will establish the necessary conditions for this type of bifurcation to occur by applying a theory based on the general center manifold theory [11].

### 3.3. Characterization of the backward bifurcation

Considering the results established in [11] based on the center manifold theory, we will now determine the existence of other equilibria and the local stability of the non-hyperbolic equilibrium, characterizing the backward bifurcation. For convenience and better guidance of the reader, we will follow the notation used in the reference [11].

The system (1) of differential equations is expressed as

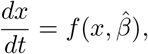

where *x* is an 8-dimensional vector with components *x* = (*x*_1_, *x*_2_, *x*_3_, *x*_4_, *x*_5_, *x*_6_, *x*_7_, *x*_8_)^*T*^. These components correspond to the variables *S* = *x*_1_, *I*_*P*_ = *x*_2_, *R*_*P*_ = *x*_3_, *S*_*P*_ = *x*_4_, *I*_*S*_ = *x*_5_, *R* = *x*_6_, *S*_*V*_ = *x*_7_ and *I*_*V*_ = *x*_8_ respectively. The parameter 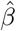 influences the dynamics of the system, and the function

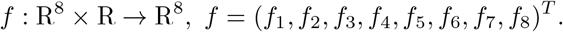

The system (1) can be rewritten as

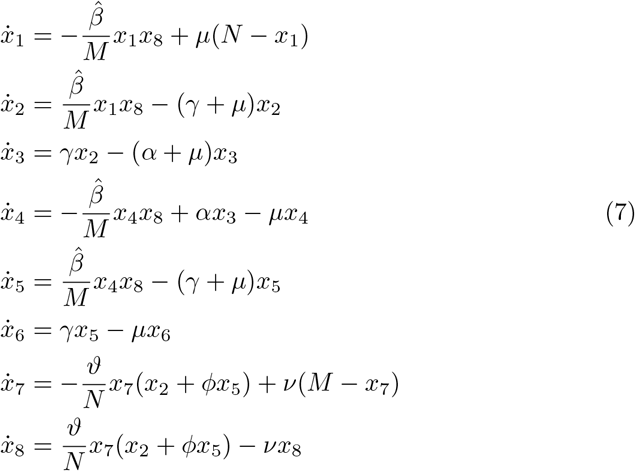

The equilibrium *x*_0_ := *E*_0_ = (*N*, 0, 0, 0, 0, 0, *M*, 0) described in the previous section is an equilibrium of the system (7) for all values of the parameter 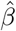. That is

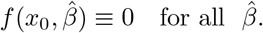

Now we will check the assumptions in [11, Theorem 4.1].

#### Assumption A1

Upon simple inspection of the Jacobian matrix *J* of the system (7 - 14) at the disease-free equilibrium *x*_0_ = (*N*, 0, 0, 0, 0, 0, *M*, 0), zero is a simple eigenvalue of *J* when 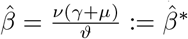.

Note that this occurs when ℛ_0_ = 1, hence the transmission rate 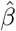 is the chosen bifurcation parameter for this analysis.

The Jacobian matrix at *x* = *x*_0_, with 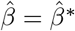, denoted by *A*, is given by

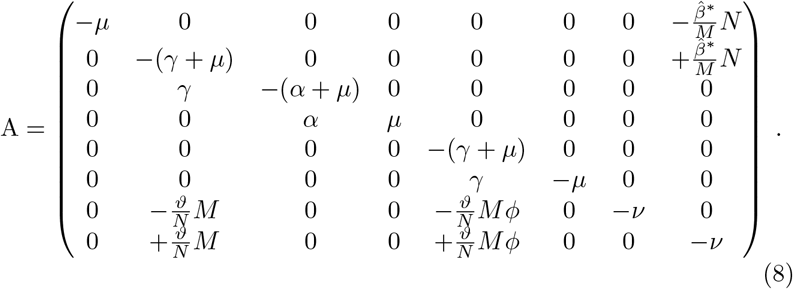

As discussed before, it satisfies the hypothesis A1 of theorem 4.1 in [11], i.e., matrix *A* has a simple zero eigenvalue, and all the other eigenvalues are negative.

#### Assumption A2

The second hypotheses required the calculation of the right and left eigenvector as follows. The matrix *A* has a right eigenvector

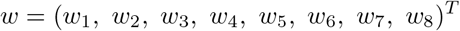

(corresponding to the zero eigenvalue) given by

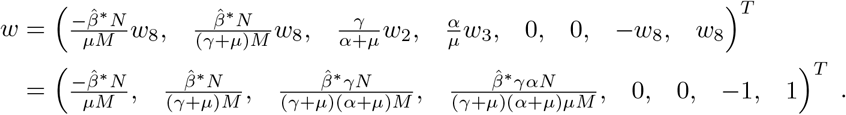

The matrix *A* has a left eigenvector *v* = (*v*_1_, *v*_2_, *v*_3_, *v*_4_, *v*_5_, *v*_6_, *v*_7_, *v*_8_) (corresponding to the zero eigenvalue) given by

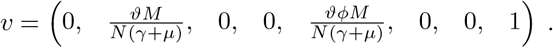

Considering the assumptions above A1 and A2 the Theorem 4.1 in [11] guaranties that the nature of the endemic equilibria near to the bifurcation point is determined by the sign of the numbers *a* and *b* given by

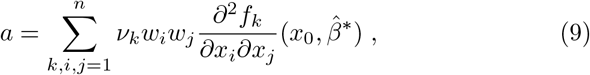

and

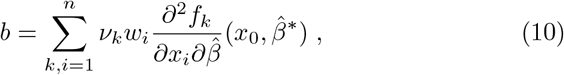

with *f*_*k*_ being the *k*^*th*^ component of *f*, and see reference [11].

#### Observation

Note that in this work 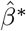 is when a zero eigenvalue occur for the matrix *A*, representing the bifurcation value *ϕ*≡ 0 in the cited reference.

Therefore, calculating these numbers we get

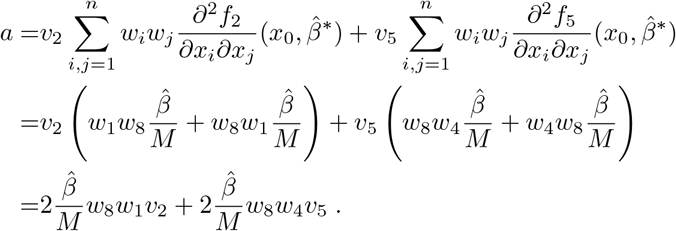

And thus,

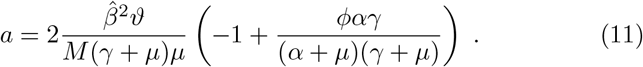

For the computation of *b*, we have

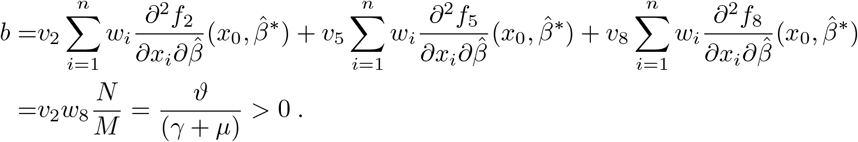

Thus, *b* is always positive. And, the sign of *a* follow these inequalities:

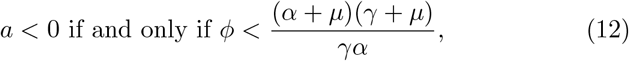

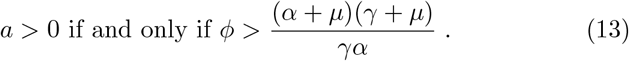

Therefore, the existence of positive equilibria, and consequently its stability, will also depend on the parameter *ϕ*. We formalized these results in Theorem 3.3 below, in accordance with the theorems in [11].

##### Theorem 3.3.

*The system exhibits a backward bifurcation at ℛ*_0_ = 1 *if, and only if*, 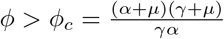.

#### Observation

In other words, the system exhibits a subcritical transcritical bifurcation at 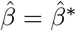 if, and only if, 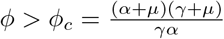. And, a supercritical transcritical bifurcation at 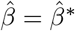 if, and only if, 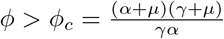.

To illustrate the analytical results of Theorem 3.3, Figure 3 (a) show for *ϕ < ϕ*_*c*_, that transcritical supercritical bifurcation occurs at ℛ_0_ = 1. For *ϕ > ϕ*_*c*_, see Figure 3 (b), subcritical transcritical bifurcation occur (backward bifurcation).

**Figure 3.**
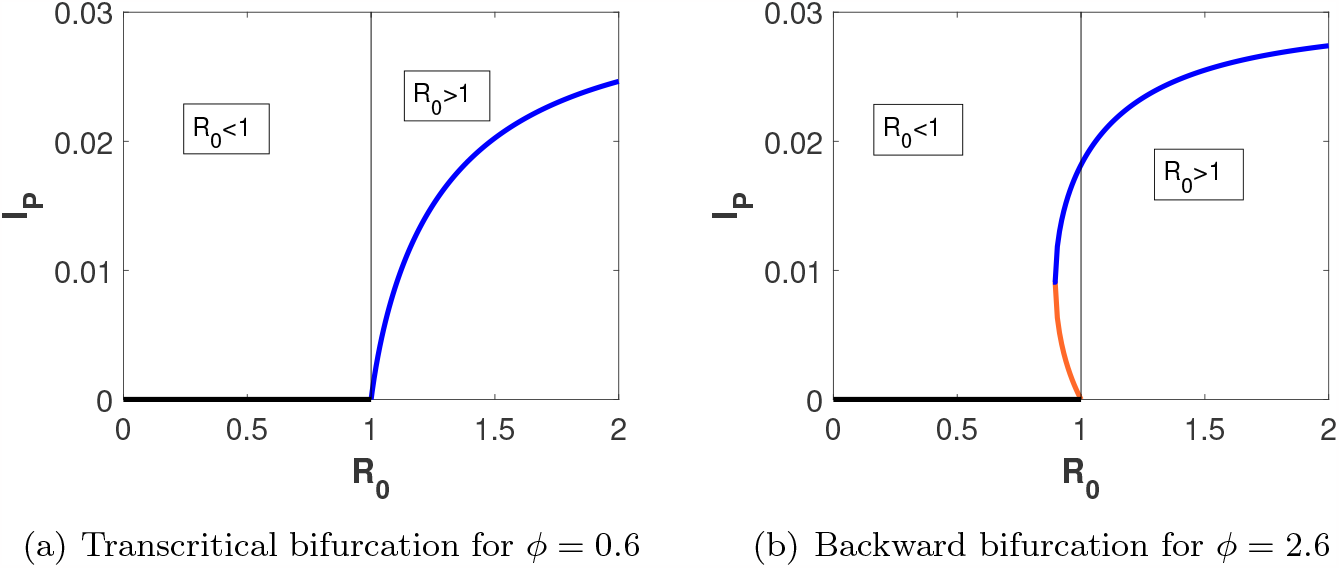
Graphical representation of the system’s equilibria (for primary infected population) when 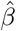is varying, for fixed parameter values shown in Table 2. The black lines represent the DFE. In a) the blue line represents the unique endemic equilibrium. In b) the blue and orange lines represent the lower and upper branch of endemic equilibria.

### 3.4. Analysis of the stability of the equilibria

The outcomes of Theorem 4.1 (refer to [11]) ensure that for *a, b >* 0, not only the equilibria exist, but their stability is also assured. Consequently, when *ϕ > ϕ*_*c*_ and 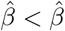, the Disease-Free Equilibrium (DFE) is locally asymptotically stable, and an additional positive unstable equilibrium, denoted as *E*_1_, emerges. However, when 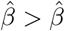, the DFE becomes unstable, resulting in the presence of a negative and locally asymptotically stable equilibrium, albeit lacking biological significance.

**Table 2:** Baseline parameter values used for simulations. The baseline values where obtained from [5, 3, 25, 28].

| Parameters | Description | Baseline values |
| --- | --- | --- |
| $\hat{\beta}$ | transmission rate from vector to humans | $52 y^{-1}$ |
| $\gamma$ | recovery rate | $52 y^{-1}$ (1 week) |
| $\alpha$ | temporary immunity rate | $52 y^{-1}$ (1 week)<br>$2 y^{-1}$ (6 months)<br>$0.5 y^{-1}$ (2 years) |
| $\phi$ | ratio of secondary infections contribution to the overall force of infection | 0.6 or 2.6 |
| $\mu$ | natural mortality and birth rates (humans) | $1/65 y^{-1}$ |
| $\vartheta$ | transmission rate from human to vector | $73 y^{-1}$ (2 $\nu$) |
| $\nu$ | natural mortality and birth rate (mosquito) | $36.5 y^{-1}$ (20 days) |
| $\eta$ | seasonal amplitude | 0.35 |
| $T$ | seasonal period | 1 y |
| Constants |  |  |
| $N$ | total human population | 100 |
| $M$ | total mosquito population | 1000 |

Conversely, for *a <* 0, *b >* 0, in instances where *ϕ < ϕ*_*c*_ and 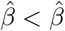, the DFE exhibits local asymptotic stability. Yet, when 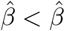, the DFE loses stability, and a formerly negative unstable equilibrium transforms into a positive and locally asymptotically stable one.

These findings are systematically outlined in the subsequent theorems.

#### Theorem 3.4.

*The disease-free equilibrium E*_0_ *of the system (1) is locally asymptotically stable if ℛ*_0_ *<* 1, *and it is unstable if ℛ*_0_ *>* 1.

*Proof*. The Jacobian matrix of the model system (1) at the disease-free steady state *E*_0_ is given by

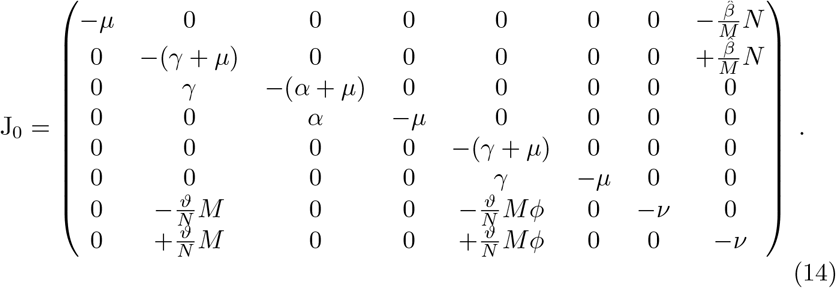

he Jacobian matrix of the model system (1) at the disease-free steady state *E*_0_ is characterized by six eigenvalues: −*μ*, − (*α* + *μ*), −*μ*, − (*γ* + *μ*), −*μ* and −*ν*. The remaining two eigenvalues are determined as the roots of the quadratic equation:

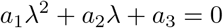

where *a*_1_ = 1 *>* 0, *a*_2_ = (*ν* + *μ* + *γ*) *>* 0, *a*_3_ = *ν*(*γ* + *μ*)(1 −ℛ_0_) *>* 0 for ℛ_0_ *<* 1. According to the Routh-Hurwitz criteria, the condition *a*_1_ = 1 *>* 0, *a*_1_*a*_2_ −*a*_3_ *>* 0 ensures that *E*_0_ will be locally asymptotically stable when ℛ_0_ *<* 1.

Additionally, in the case where ℛ_0_ *>* 1 there is one positive eigenvalue *λ*_*m*_ while the others remain negative. The analytical expression for *λ*_*m*_ is given by:

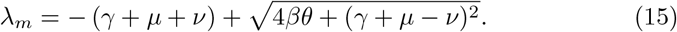

At the critical point

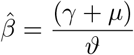

this eigenvalue undergoes a change in sign as 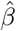 increases, transitioning from negative to positive. This sign change is supported by the positive derivative for 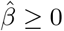:

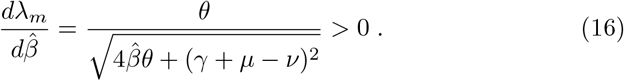

#### Theorem 3.5.

*The endemic equilibrium E*_1_ *of the system (1) is locally asymptotically stable if ℛ*_0_ *>* 1 *and (ϕ < ϕ*_*c*_).

#### Observation

Although the analytical proof of this theorem can be obtained in its closed form, it involves dependencies on three parameters, leading to laborious computations without significant additional insight (see more in Appendix 6.2). Consequently, we direct the reader to the numerical findings that represent the results of this theorem.

## 4. Numerical simulations and bifurcations structures

To facilitate a direct comparison with the results in [34], where the host-host SIRSIR model is discussed, we introduce the definition 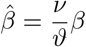 (as proposed in [26]). Here, *β* is defined in [34], and the basic reproduction number is denoted by 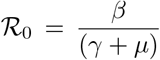. Table 2 lists the parameter values set as baseline for the numerical simulations.

### 4.1. Stability of endemic equilibria

In order to illustrate the stability results for the endemic equilibria, the real part of the largest eigenvalues is plotted in Figure 4. Here, the transmission rate *β* is varied for two values of *ϕ* (*ϕ* = 0.6 *<* 1 and *ϕ* = 2.6 *>* 1), considering three scenarios for the immunity period, *α*: short (*α* = 52*y*^−1^ = 1 week), intermediate (*α* = 2*y*^−1^ = 6 months), and long (*α* = 2*y*^−1^ = 2 years), as evaluated also in [34]. The transcritical bifurcation, i.e., ℛ_0_ = 1, occurs for the parameter values fixed in Table 2 at *β*≈ 52 *y*^−1^, or equivalently, 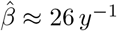.

**Figure 4.**
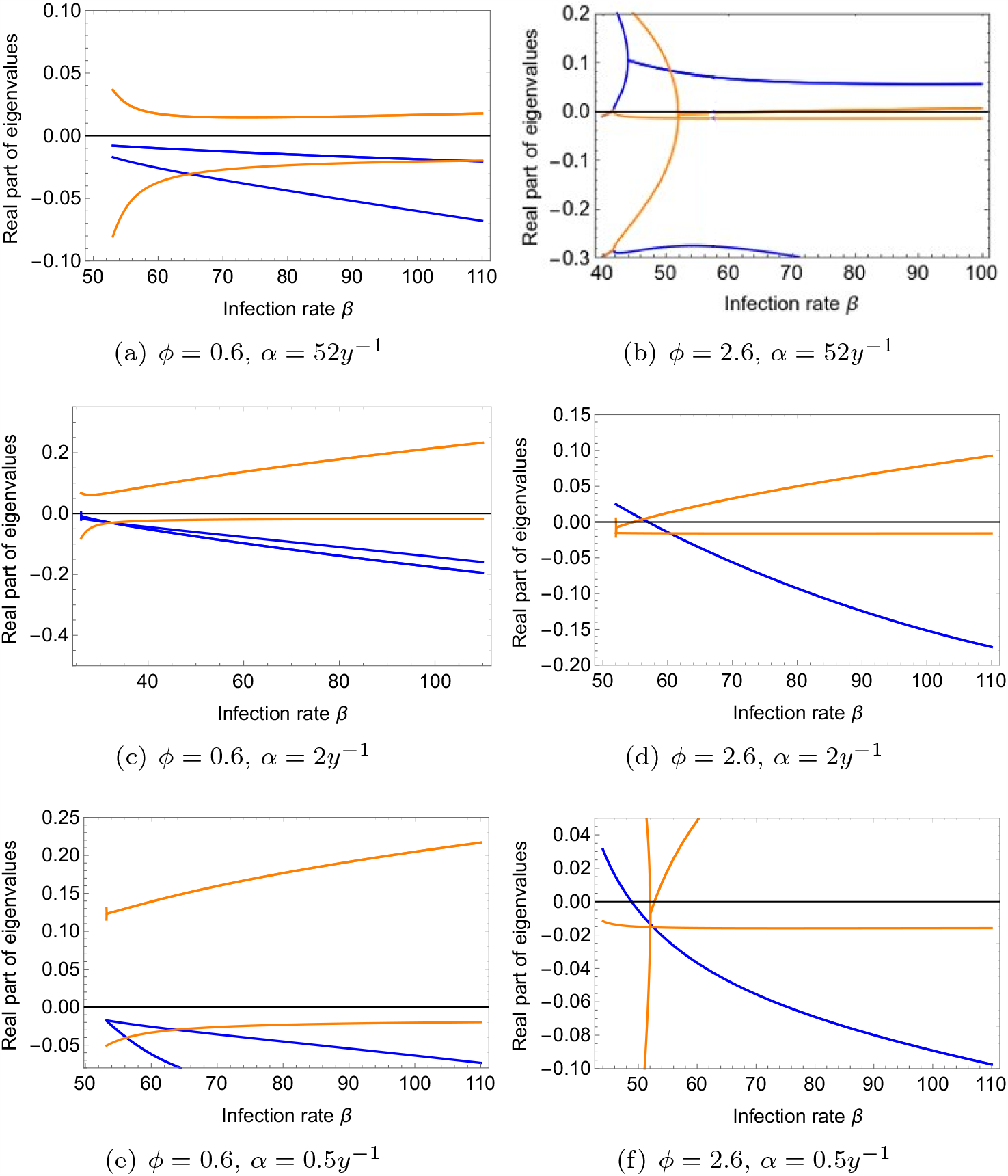
Real part of the eigenvalues of the endemic equilibria when 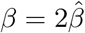 is varying, for fixed parameter values shown in Table 2. The blue line represents the real part of the largest eigenvalue of the positive (upper branch) equilibria. The orange line represents the real part of the largest eigenvalue of the lower branch equilibria.

According to the analytical results of the theorems described in the previous section, for a lower period of immunity (*α* = 52*y*^−1^) and *ϕ* = 0.6 *< ϕ*_*c*_, the system exhibits a unique positive stable endemic equilibrium (see the blue line in Fig. 4 (a)). However, for *ϕ* = 2.6 *> ϕ*_*c*_, the positive equilibrium is asymptotically unstable (see the blue line in Fig. 4 (b)), and a periodic orbit is stable (not shown), leading to a periodic solution for this set of parameter values.

For an intermediate period of immunity (*α* = 2*y*^−1^), when *ϕ* = 0.6 *< ϕ*_*c*_, the system exhibits a unique positive stable endemic equilibrium. When 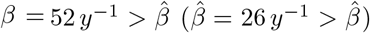, the positive equilibrium is asymptotically stable (see the blue line in Fig. 4 (c)). When 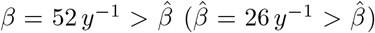, the lower branch becomes a negative equilibrium and locally asymptotically stable, only for a small range of parameter values, becoming unstable again (see the orange line in Fig. 4 (d)), and the upper branch (positive equilibrium) becomes asymptotically stable (see the blue line in Fig. 4 (d)).

Finally, for longer periods of immunity (*α* = 0.5*y*^−1^) and *ϕ* = 0.6 *< ϕ*_*c*_, similar results are observed compared to an intermediate period of immunity, exhibiting a unique positive stable endemic equilibrium (see the blue line in Fig. 4 (e)). On the other hand, stability of the endemic equilibrium is observed after the transcritical bifurcation, i.e., after ℛ_0_ *>* 1. Also, bi-stability occurs before the transcritical bifurcation, i.e., for small parameter value regions, the DFE and the endemic equilibria are stable (see the blue line in Fig. 4 (f)). Bi-stability occurs for 2· 24.5 *y*^−1^ *< β* = 2^*β <* 2 ·26 *y*^−1^ for *α* = 0.5*y*^−1^ and *ϕ* = 2.6.

### 4.2. Two dimensional bifurcation diagrams for the parameters 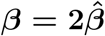 and *ϕ*

In the previous sections, we observed that the value of the parameter *ϕ* defines whether there is a subcritical or supercritical transcritical bifurcation, i.e., whether the DFE is the only positive equilibrium before ℛ_0_ = 1. Therefore, the two-dimensional analysis of these two main parameters, the transmission rate *β* and the enhancement factor *ϕ*, will illustrate the bifurcation structures present in this 9-dimensional model.

Figures 5 (a), 5 (b), and 5 (c) show the 2-parameter bifurcation diagrams for the SIRSIR-UV model, corresponding to each of the three scenarios of immunity periods, *α* = 52*y*^−1^, *α* = 2*y*^−1^, and *α* = 0.5*y*^−1^, respectively. The bifurcation parameter is *ϕ*, with 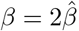, while all other parameters are set as described in Table 2.

**Figure 5.**
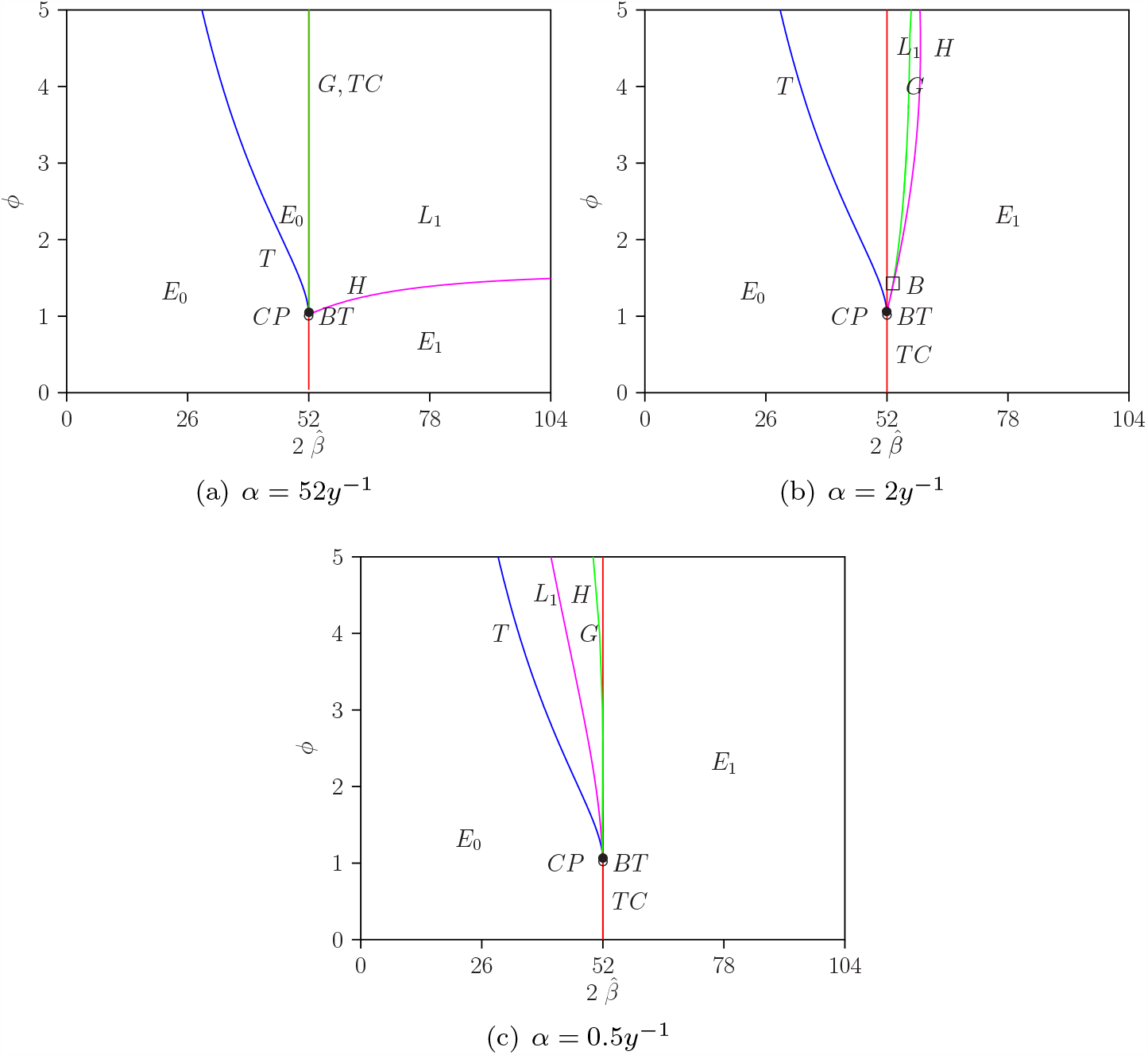
Two-parameter bifurcation diagram for 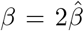 and *ϕ* (and *η* = 0) for the SIRSIR-UV model. (a) for *α* = 52*y*^−1^, (b) for *α* = 2*y*^−1^, and (c) for *α* = 0.5*y*^−1^.

The Tangent (*T*) bifurcation curve is represented by the color blue, Trans-critical (*TC*) by red, Hopf (*H*) by magenta, and global homoclinic (G) by green. Cusp bifurcation (*CP*), Bogdanov-Takens point (*BT*), and Bautin (B) are also depicted in the figures. *E*_0_ is represented in the region where the DFE is stable, *E*_1_ where the endemic equilibrium is stable, and *L*_1_ where a stable limit cycle appears as a solution of the system. For a more detailed explanation of each bifurcation structure, the reader is referred to [22].

In Figure 5, it is noteworthy that, starting from the codimension-2 bifurcations, the codimension-1 curves divide the parameter space into regions with different long-term dynamics, such as the disease-free equilibrium (*E*_0_), the endemic equilibrium (*E*_1_), and the limit cycles (*L*_1_).

#### 4.2.1. One dimensional bifurcation diagram for the parameter *ϕ*

Combining the analysis of stability of endemic equilibria in Fig. 4 (b) – where it is demonstrated to be unstable – with the observation of a Hopf bifurcation curve in the two-dimensional bifurcation diagram in Figure 5 (a), we can deduce that a Hopf bifurcation occurs for a specific fixed value of *β*. This implies that for a given ℛ_0_, the equilibrium is stable before reaching a critical value of *ϕ*, periodic oscillations are observed at the critical value *ϕ*_*c*_, and after *ϕ*_*c*_, the equilibrium becomes unstable, and the limit cycle becomes stable.

To provide a comprehensive overview, a bifurcation diagram (depicting the maximum value for (*I*_*P*_) in blue and the minimum value in red) is presented in Figure 6 (a), (b), and (c), utilizing *ϕ* as the bifurcation parameter while keeping 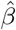 fixed. Figure 6 (a) illustrates that the Hopf bifurcation occurs at *ϕ* = 1.45 for 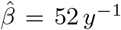, exclusively for a short period of immunity (*α* = 52 *y*^−1^). For intermediate and longer periods, the endemic equilibrium remains stable after the transcritical bifurcation.

**Figure 6.**
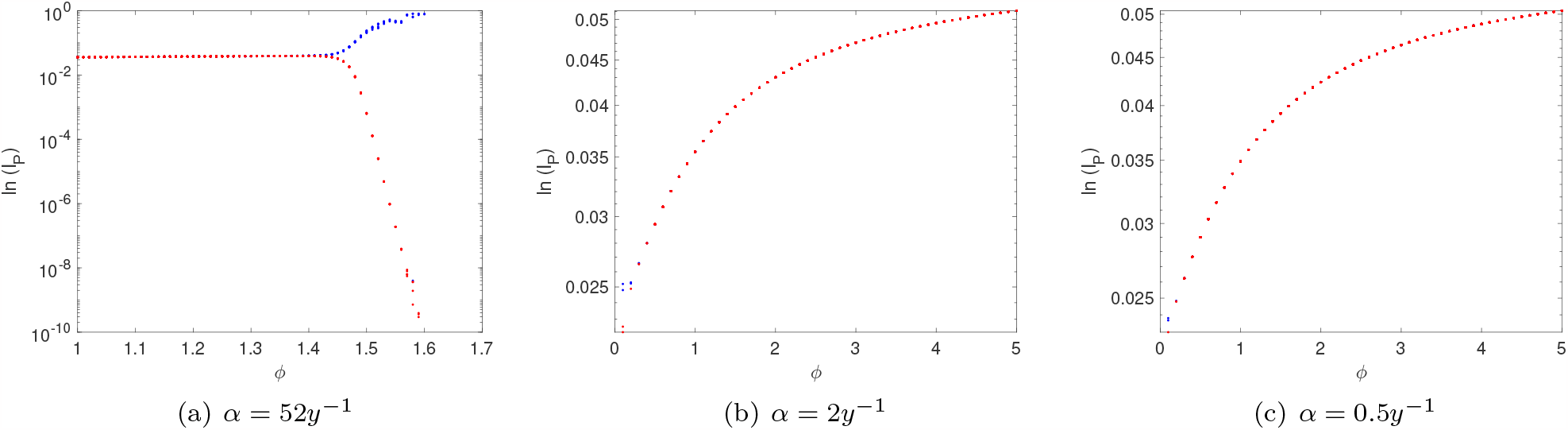
Bifurcation diagram showing minimal (in red) and maximal (in blue) values for total infection (*I*_*P*_) with respect to the parameter *ϕ* for *β* = 52 *y*^−1^, *γ* = 52*y*^−1^, *η* = 0, and *ϕ* ranging from 0 to 5. Panels depict different scenarios for the immunity period *α*: (a) for *α* = 52*y*^−1^, (b) for *α* = 2*y*^−1^, and (c) for *α* = 0.5*y*^−1^.

#### 4.2.2. One dimensional bifurcation diagram for the parameter 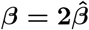

In the continuation we highlight 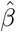 as a bifurcation parameter and fixing the parameter *ϕ*. In Fig. 7 one dimensional diagram for different period of immunity is shown, that is, for small, intermediate and longer period of immunity, *α* = 52, 2 and 0.5*y*^−1^, respectively. The parameter *ϕ* is fixed to 2.6.

**Figure 7.**
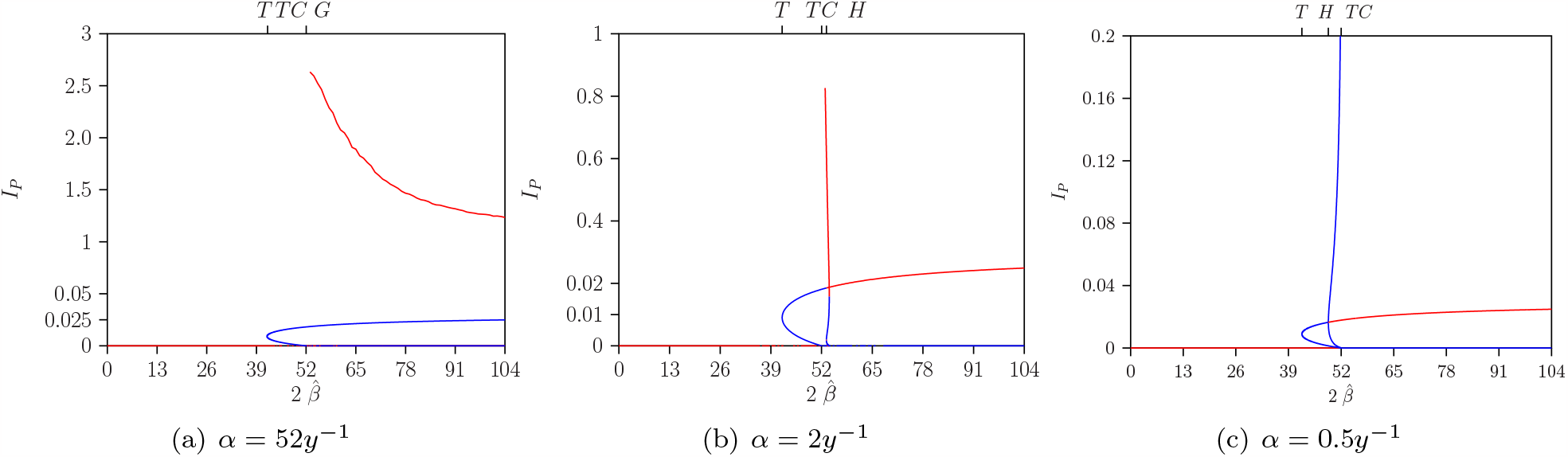
Bifurcation diagram for the parameter 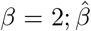 with *ϕ* = 2.6, *η* = 0, and other parameters described in the table. (a) for *α* = 52*y*^−1^, (b) for *α* = 2*y*^−1^, and (c) for *α* = 0.5*y*^−1^. Color red represents stable equilibria, while the color blue represents unstable ones.

Note that in Figure 7 for a fixed *ϕ* = 2.6, the *T, TC, G*, and *H* bifurcation points are observed, as already depicted in Figure 5. For a small temporary period, *α* = 52*y*^−1^, after the transcritical bifurcation, the endemic equilibrium is already unstable, and a global homoclinic point establishes the dynamics of the system, leading to a stable limit cycle (see Fig. 8).

**Figure 8.**
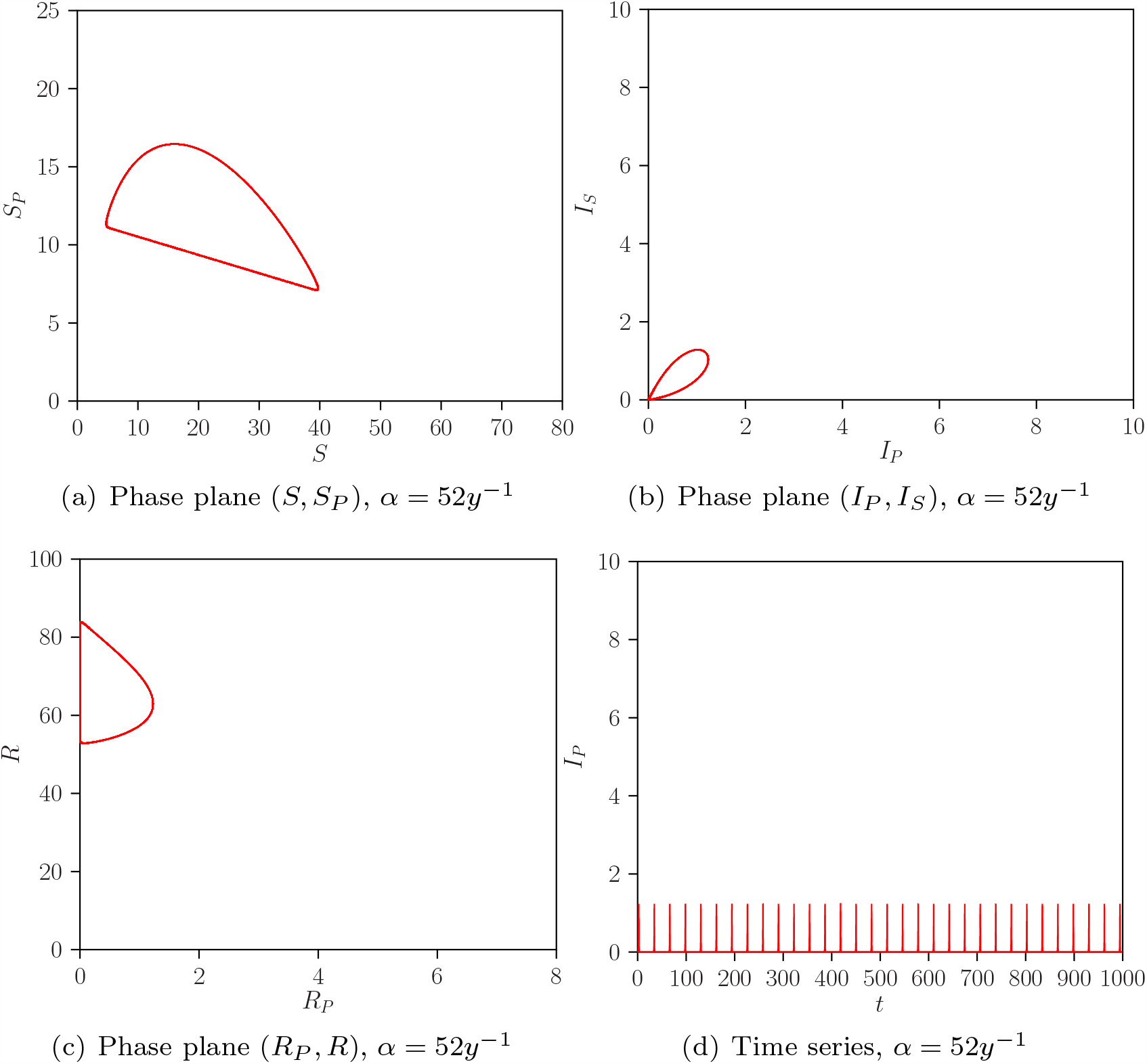
Projection phase plane (a), (b), and (c) and time evolution (d) plots for the autonomous SIRSIR-UV model (*η* = 0) with *α* = 52*y*^−1^, *ϕ* = 2.6, and 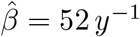.

However, for intermediate and larger periods, i.e., *α* = 2 and 0.5*y*^−1^, a unique positive stable equilibrium is observed after the Hopf bifurcation. The difference between these two scenarios is that, while for the intermediate period, the Hopf bifurcation occurs after the transcritical bifurcation, for the larger period, it occurs before the transcritical bifurcation. This once again shows that for a small window, bi-stability occurs between the DFE and the endemic equilibrium, for a larger period (see Fig. 7 c).

## 5. Modelling mosquito abundance changes due to seasonality factors

In this section, we incorporate an external force into the model to capture ecological factors influencing mosquito abundance, specifically, the seasonality of the mosquito population influenced by climate factors like temperature or rainfall. The mosquito population, represented by *M*, is modeled as a constant population that undergoes seasonal variations described explicitly by a cosine function, as follows:

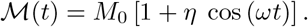

In the equation above, *M*_0_ represents the mean mosquito population, and *η* describes the magnitude of the seasonal amplitude, where *ω* = 2*π/T*. The frequency parameter models yearly variations in the mosquito population. Incorporating this external force into the model, it’s important to note that the system of ordinary differential equations (1) transforms into a non-autonomous ODE system.

### 5.1. A closer analysis on the theory of the non-autonomous system

In the first theoretical analysis of the non-autonomous system with seasonality, it’s noteworthy that, concerning the time dependence, the host and vector models are decoupled. This decoupling can be used in the analysis of the disease-free state, where *S*(*t*) = *N, I*_*P*_ (*t*) = 0, *R*_*P*_ (*t*) = 0, *S*_*P*_ (*t*) = 0, *I*_*S*_(*t*) = 0, *R*(*t*) = 0 for all *t*.

In this disease-free state, the host-vector model can be expressed as follows:

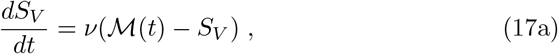

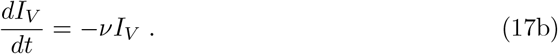

The solution thus yields the limit cycle *L*_0_ described by *S*_*V*_ = ℳ (*t*) and *I*_*V*_ = 0. Numerical analysis results indicate that this holds for 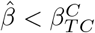, which is generally not the same as *β*_*T C*_. It is a transcritical bifurcation of a limit cycle, and an invasion criterion for this limit cycle is similar to how 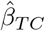 was for the autonomous disease-free equilibrium case. In general, only non-stationary endemic solutions exist. There are solutions with the same period as the forcing but also forcing-induced cyclic, quasi-stationary, and chaotic solutions.

Numerical results are shown in Fig. 9 for parameter values in Table 2, where the dominant absolute value of the Floquet multiplier is shown depending on the parameter 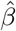.

**Figure 9.**
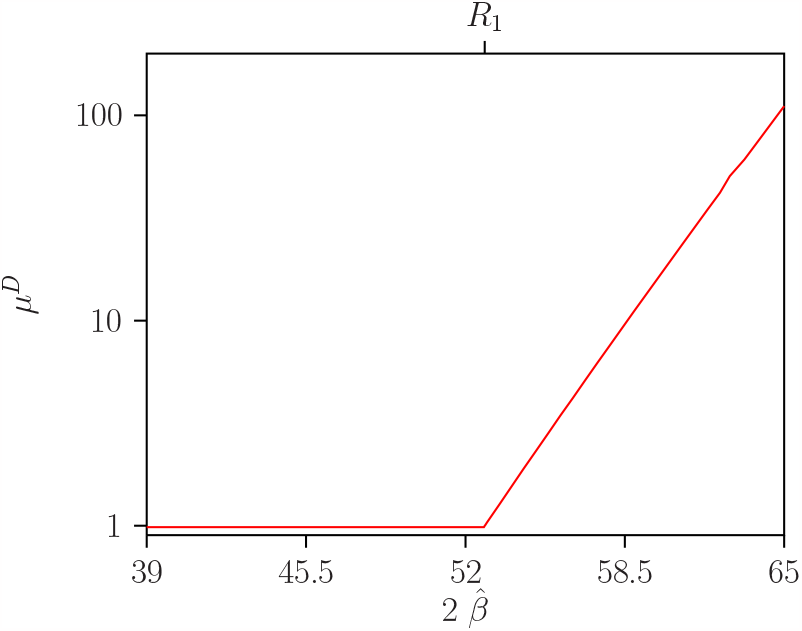
Dominant multipliers *μ*^*D*^ as function of 2 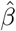 for disease-free cycle *L*_0_ where *γ* = 52*y*^−1^, *η* = 0.35, *ϕ* = 2.6, *α* = 52*y*^−1^. The resonance 1:1 bifurcation point *R*_1_ occurs at 2 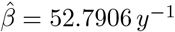.

The point *R*_1_ (shown in Fig. 9) marks a co-dimension 2 1:1-resonance point where two dominant multipliers *μ*^*D*^ are equal to 1 [22, 21]. It occurs at 2 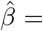52.7906 *y*^−1^, hence slightly (depending on *η*) larger than 2 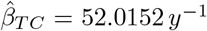. In this case, it is also a transcritical bifurcation for the limit cycle *L*_0_. For parameter values before this point, the cycle *L*_0_ is stable because *μ*^*D*^ *<* 1, and above the point where *μ*^*D*^ *>* 1, a non-cyclic endemic solution prevails.

### 5.2. Numerical bifurcation analysis of non-autonomous model

The numerical experiments in the following sections used the baseline values stated in Table 1, unless stated otherwise. For the seasonal forcing function parameters, we used *η* = 0.35 and a period *T* = 1 *y*.

#### 5.2.1. Two dimensional bifurcation diagrams for 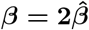 and *ϕ* with *η* = 0.35

The two-dimensional analysis of the two main parameters, the transmission rate 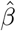, and the enhancement factor *ϕ*, provides a visualization of the bifurcation structures in this non-autonomous model. Figure 10 shows the two-parameter bifurcation diagrams for the host-vector SIRSIR-UV non-autonomous model, considering different scenarios of immunity periods *α* = 52*y*^−1^, *α* = 2*y*^−1^, and *α* = 0.5*y*^−1^, with all other parameters set to their baseline values.

**Figure 10.**
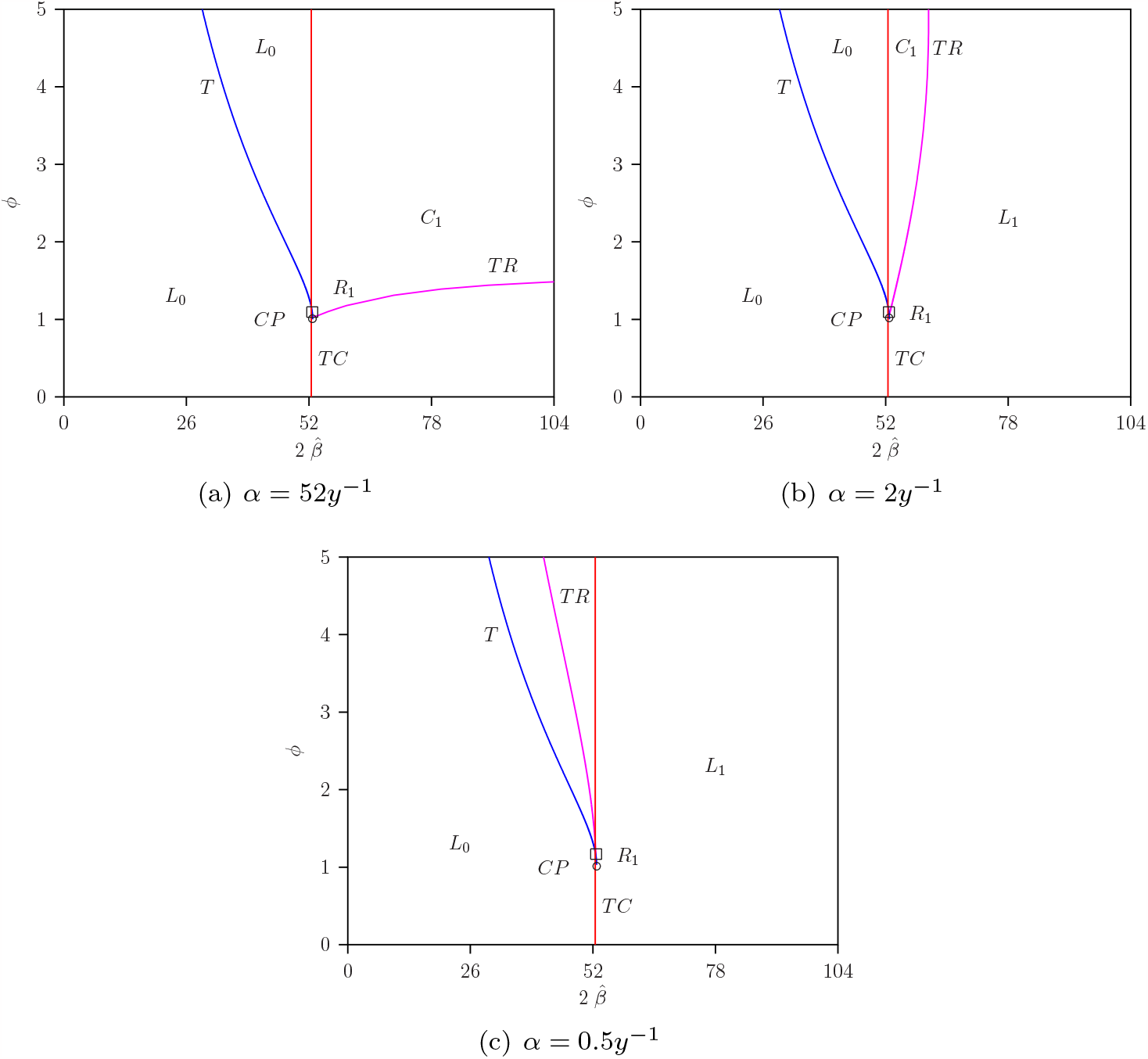
Two-parameter bifurcation diagram sowing the relationship between 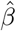 and *ϕ* for the non-autonomous model (where *η* = 0.35). In each diagram, the scenarios for different values of *α* are considered: (a) *α* = 52*y*^−1^, (b) *α* = 2*y*^−1^, and (c) *α* = 0.5*y*^−1^. The vertical line, where 2*β* = 52.7906 *y*^−1^, represents the *R*_1_ curve and serves as the transcritical curve for the disease-free limit cycle in all diagrams.

In the figure 10, the Tangent (*T*) bifurcation curve is represented by the color blue, Transcritical (*TC*) by red, torus (*TR*) by magenta. Cusp bifurcation (*CP*) and co-dimension 2 1:1-resonance points (*R*_1_) are also depicted. Regions denoted as *L*_0_ represent where the disease-free limit cycle is stable, while *L*_1_ denotes where the endemic periodic solution is stable, and *C*_1_ denotes a chaotic region.

The point *R*_1_ signifies a co-dimension 2 1:1-resonance point on fold or torus bifurcation curves, providing a more detailed analysis, as shown in Fig. 10.

The parameter region is completely determined and divided into regions where the disease-free limit cycle is stable, the endemic limit cycle is stable, or a chaotic regime resulting from a torus bifurcation.

Comparing the bifurcation diagrams between the non-seasonal model (*η* = 0) in Figure 5 (a) and the seasonal model (*η >* 0) with *α* = 52*y*^−1^ in Figure 10 (a) reveals that the introduction of periodic seasonal forcing always implies chaos for 2*β >* 52.7906 *y*^−1^. On the other hand, for *α* = 2*y*^−1^, in the scenario without periodic forcing (*η* = 0), there is a homoclinic bifurcation at the transcritical bifurcation point, resulting in a periodic limit cycle (Figure 5). However, when adding periodic seasonal forcing, it leads to a stable limit cycle for 2*β >* 52.1538 *y*^−1^ (Figure 10 (b)). For *α* = 2*y*^−1^, Figure 10 (b) shows that these conclusions do not hold universally, and the phenomenon depends on the parameter *ϕ*.

Moreover, the torus bifurcation curve does not occur in the non-autonomous system at the same parameter value as the Hopf bifurcation in the autonomous case, except when *η*→ 0. In parameter regions where the limit cycle is stable, chaos is not possible. When the limit cycle is unstable, it can exhibit dynamics on a torus or become chaotic.

#### 5.2.2. One dimensional bifurcation diagram for ϕ and Lyapunov exponent analysis with η = 0.35

The numerical analysis of the non-autonomous system is presented below in a bifurcation diagram, where the enhancement factor *ϕ* is taken as a bifurcation parameter (see Fig. 11 (a), (c), and (e)). As a complement to this analysis, the respective Lyapunov exponents are shown in Figure 11 (b), (d), and (f).

**Figure 11.**
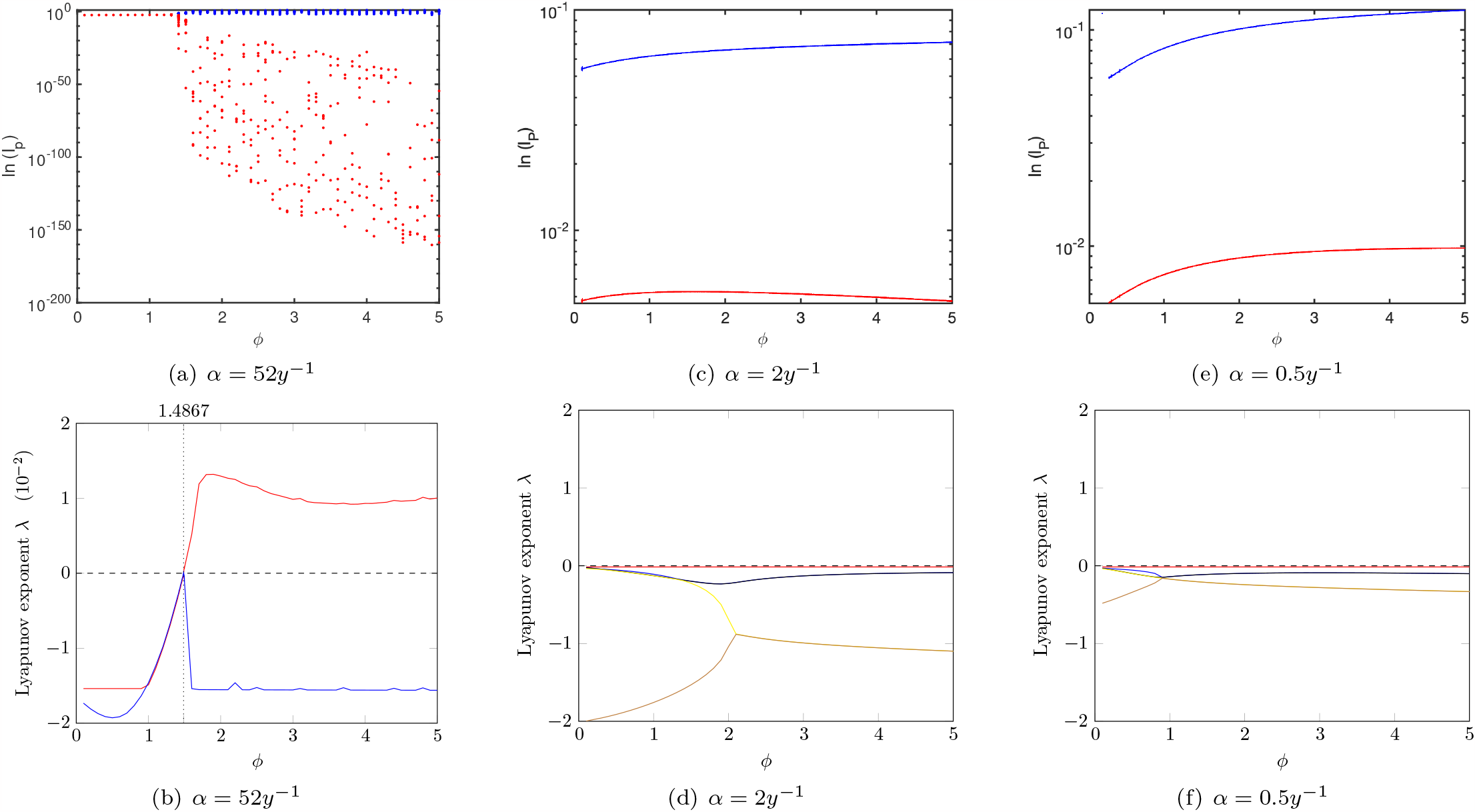
In the seasonal SIRSIR-UV model, the bifurcation diagram (upper panel) and Lyapunov spectra (lower panel) depict the largest Lyapunov exponents across various immunity periods *α*, influenced by the enhancement factor *ϕ*. Panels (a) and (b) represent a short immunity period (*α* = 52*y*^−1^), (c) and (d) correspond to an intermediate immunity period (*α* = 2*y*^−1^), while (e) and (f) illustrate a long immunity period (*α* = 0.5*y*^−1^). In (b), a vertical dotted line shows the critical *ϕ* value where the dominant Lyapunov exponent becomes positive, indicating the onset of chaotic behavior immediately after the torus bifurcation point.

While the non-seasonal system exhibits only steady states, the introduction of seasonality to the system for intermediate and longer periods of immunity (*α* = 2*y*^−1^ (Fig. 6 (c)) and *α* = 0.5*y*^−1^ (Fig. 6 (e)), respectively, results in periodic oscillations (see Fig. 11 (c) and (e)).

For a short period of immunity (*α* = 52*y*^−1^), the model without seasonality undergoes a Hopf bifurcation (Fig. 6 (a)). However, when seasonality is added to the system, chaotic behavior emerges (Fig. 11(a)), as evidenced by the presence of positive Lyapunov exponents shown in Fig. 11 (b).

The route to chaos is induced by a torus bifurcation for *α* = 52*y*^−1^, as detailed in Fig. 12. With all other parameters set to their baseline values, variations in the parameter *ϕ* result in a stable limit cycle below the torus bifurcation *TR* and chaos above it. The limit cycle becomes unstable, and the forcing leads to chaos.

**Figure 12.**
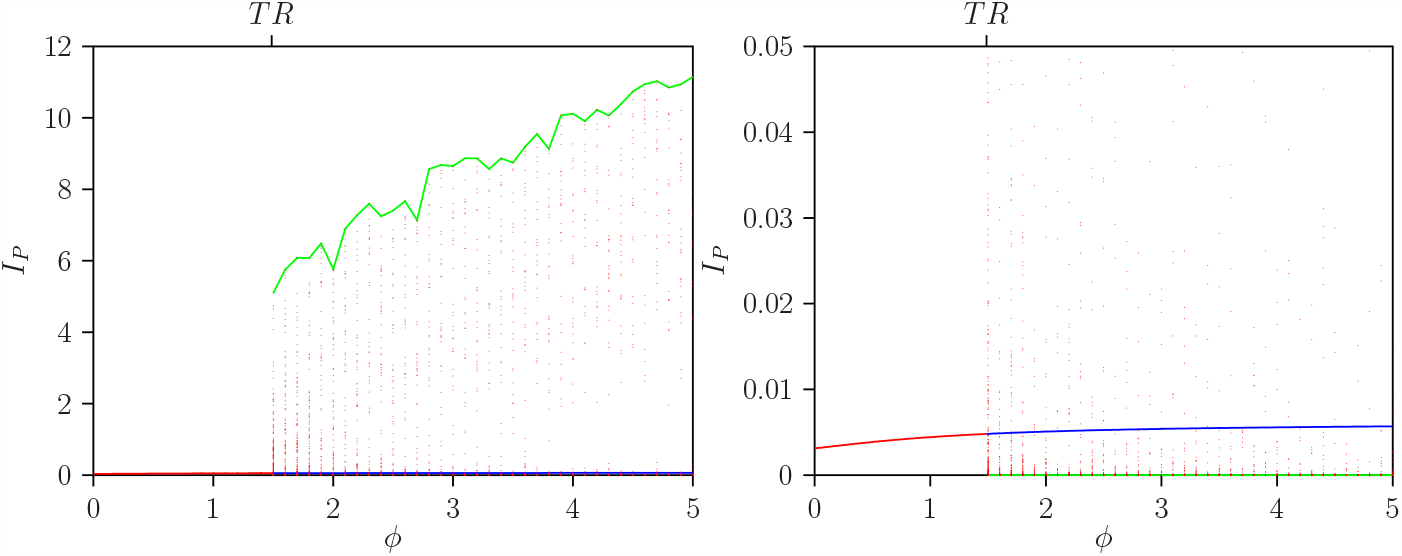
Values of *I*_*P*_ for various *ϕ* values in the non-autonomous model (*η* = 0.35) achieved after the transient period. For a short temporary immunity (*α* = 52*y*^−1^) and fixed parameters detailed in Table 2, the left figure displays global maxima, and the right one shows minima. Red dots represent local extremes, while green and blue lines denote global extremes for the state variable *I*_*P*_.

Figure 12 illustrates the minimum and maximum values for *I*_*P*_ as the parameter *ϕ* varies in the non-autonomous model. Note that the global minima for each *ϕ* value are very close to zero. This emphasizes that the results for the maxima in the top-left diagram are more discriminating for detecting chaos through simulation over time after a transient period in the chaotic region above *TR*, the global maxima (solid green line) and local (dotted red points). Maxima are shown on the left, and minima are shown on the right.

Together with Figure 11 (b), one can confirm that the Lyapunov exponents as a function of *ϕ* indicate that below the *TR* bifurcation point at *ϕ* = 1.48674, there are only stable limit cycles, and above *TR*, chaotic behavior is observed.

#### 5.2.3. One dimensional bifurcation diagram for 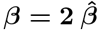 with η = 0.35

Continuing our analysis, we focus on 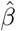 as a bifurcation parameter while fixing the parameter *ϕ* in the non-autonomous model. In Fig. 13, a one-dimensional diagram is presented for different periods of immunity: small (*α* = 52*y*^−1^), intermediate (*α* = 2*y*^−1^), and longer (*α* = 0.5*y*^−1^), while the parameter *ϕ* is held constant at 2.6.

**Figure 13.**
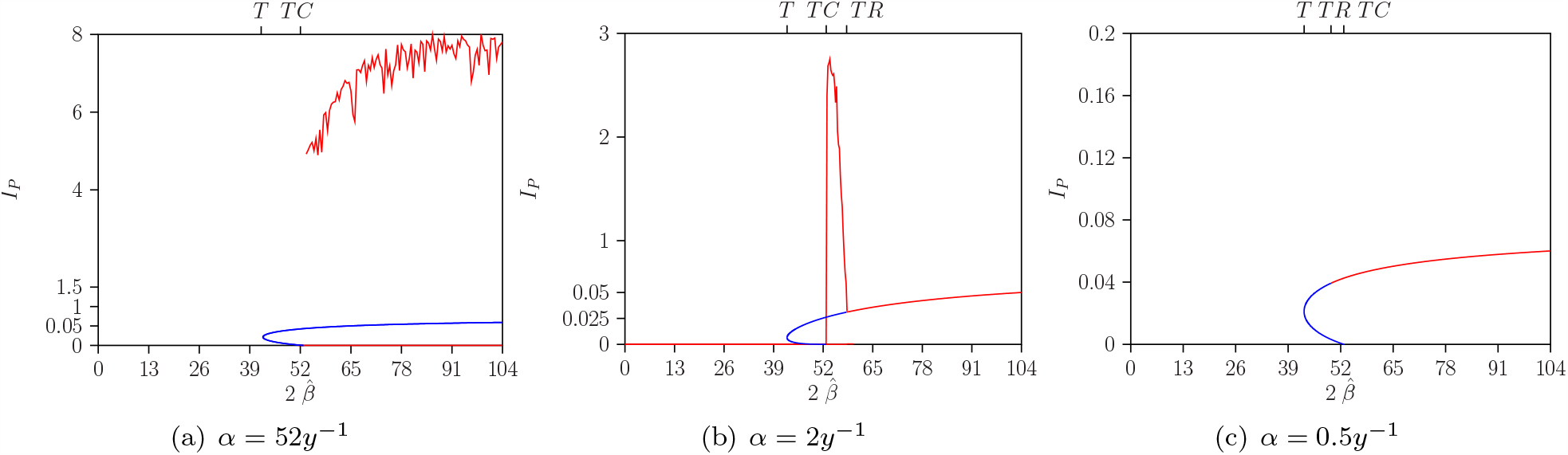
Bifurcation diagram for the non-autonomous model and parameters values are 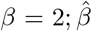 with *ϕ* = 2.6, *η* = 0.35, and other parameters described in the table. (a) for *α* = 52*y*^−1^, (b) for *α* = 2*y*^−1^, and (c) for *α* = 0.5*y*^−1^.

Figure 13 illustrates a one-parameter bifurcation diagram 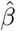 for the non-autonomous model (where *η* = 0.35). In panel (c) for *α* = 0.5*y*^−1^, the torus bifurcation *TR* occurs at 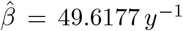, preceding *TC*. The endemic stationary dynamics are stable and unstable in parameter ranges below the torus bifurcation. In panel (b) for *α* = 2*y*^−1^, the *TR* occurs at 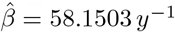, after *TC*. The endemic stationary dynamics are stable and unstable in parameter ranges below the *TR* bifurcation. In panel (a) for *α* = 52*y*^−1^, the *TR* is not visible since, for *ϕ* = 2.6 in Figure 10, it is above the point where the torus bifurcation curve *TR* intersects with the vertical borderline at 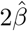. The endemic stationary dynamics are unstable in the entire parameter range above the global homoclinic bifurcation *G*. Numerical simulations show chaotic dynamics in this region (see Figure 14).

**Figure 14.**
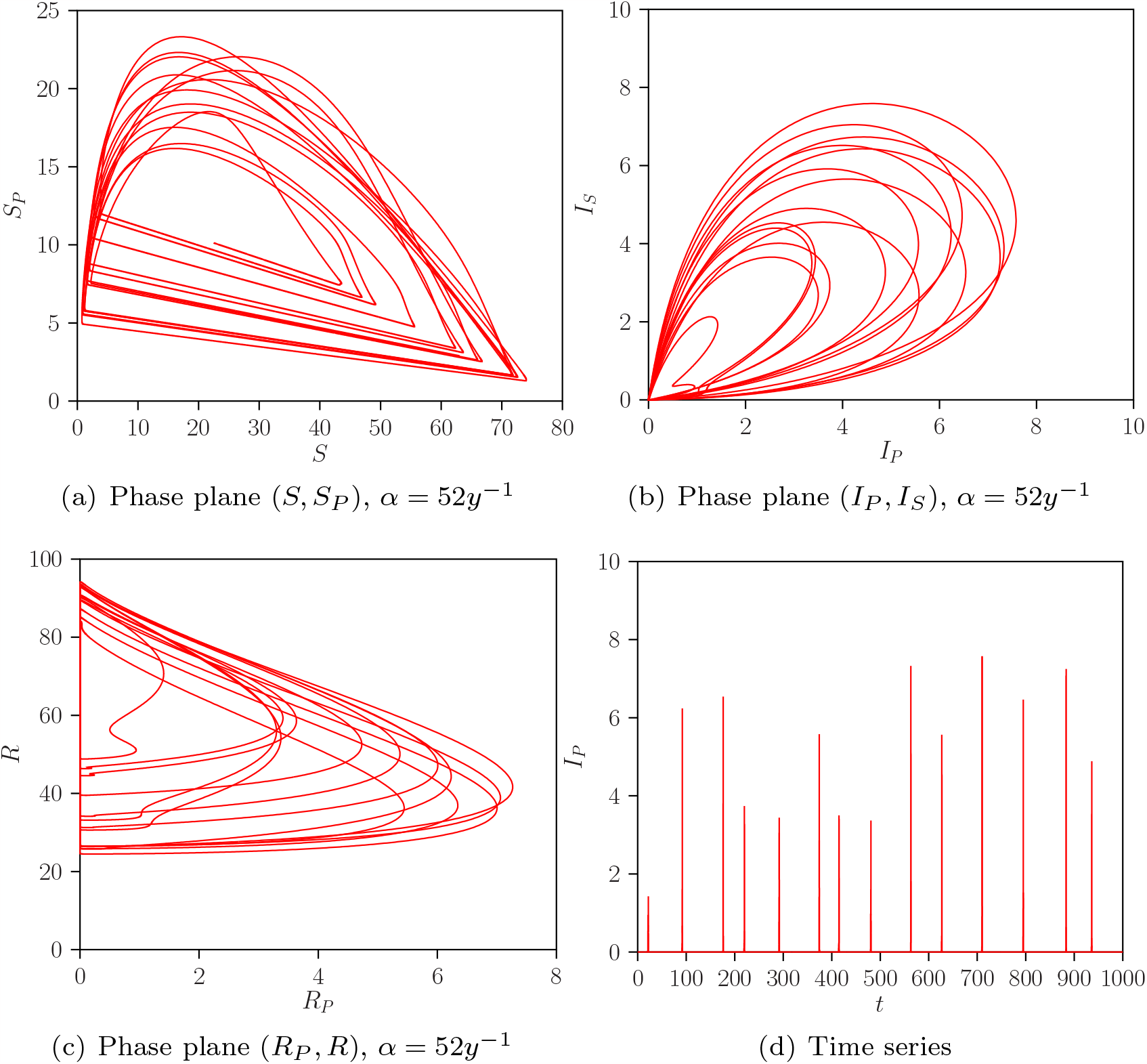
Projection phase space (a), (b) and (c) and time evolution (d) plots for non-autonomous SIRSIR-UV model, where *η* = 0.35) *α* = 52*y*^−1^. *ϕ* = 2.6, 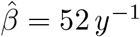.

#### 5.2.4. Route to Chaos: phase-plane plots from stable limit cycle to chaos via torus bifurcation

The transition from a stable limit cycle to chaos via torus bifurcation is detailed in Figure 15. This figure illustrates the evolution from a stable limit cycle to chaotic behavior as the parameter *ϕ* varies in the range *ϕ* ∈ [1.43, 1.5]. The *TR*-point at *ϕ* = 1.48674 is calculated using the AUTO software for continuation and bifurcation problems [14, 13]. For *ϕ* values below the *TR* point, a stable limit cycle with a negative dominant Lyapunov exponent is observed. The shape of the limit cycle becomes more complex as *ϕ* approaches the *TR* value (see Figure 15 (a-c)). Above the *TR* point, chaotic behavior is present, as indicated by the positive dominant Lyapunov exponent (see Figure 15 (d-f)).

**Figure 15.**
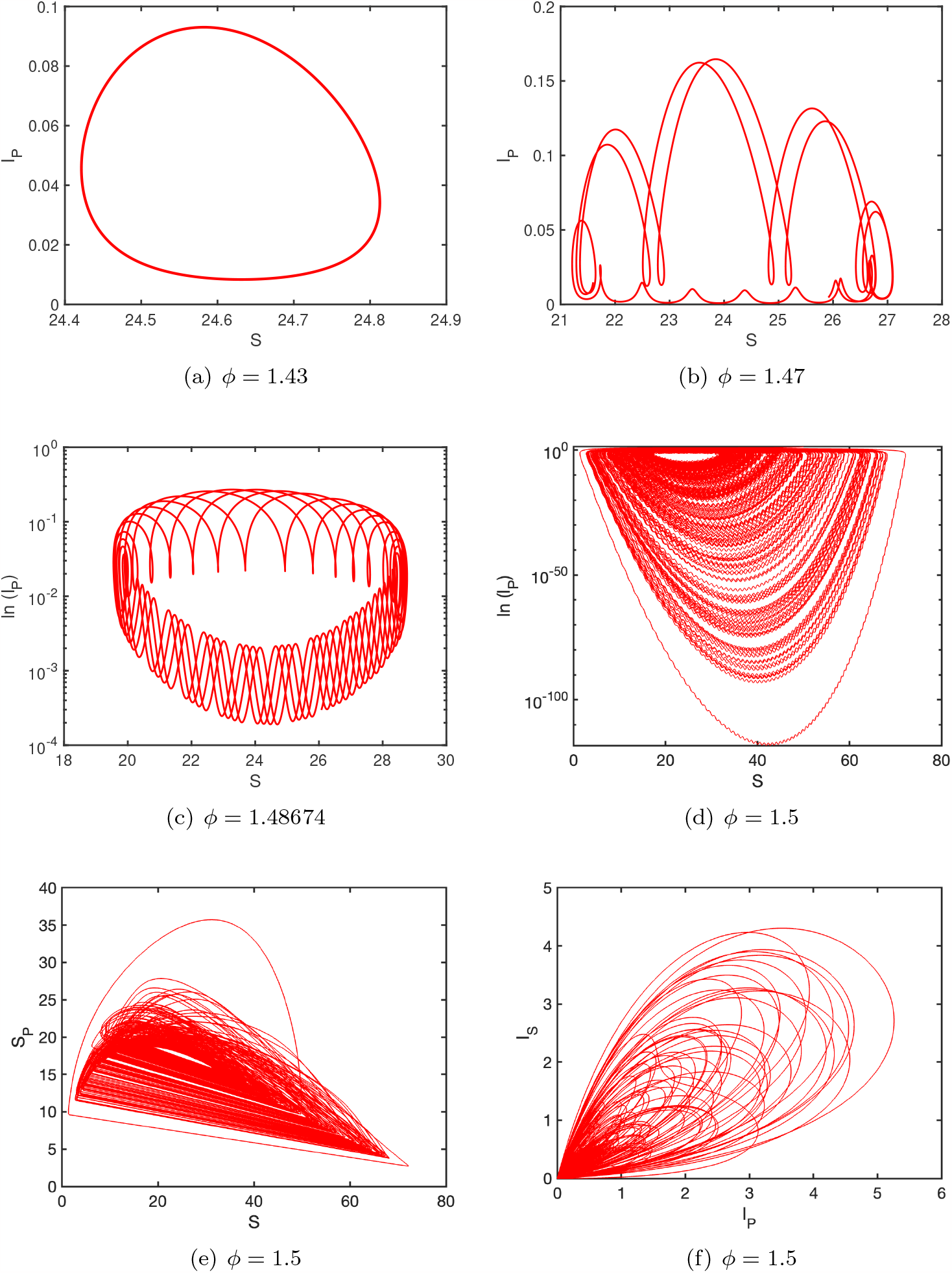
Phase space for different values of *ϕ*, fixing *β* = 52 *y*^−1^, *γ* = 52*y*^−1^, *α* = 52*y*^−1^, *η* = 0.35.

In Figure 16, the dynamics for *ϕ* = 1.46 and *ϕ* = 1.47 are combined, illustrating the transformation of the limit cycle into a torus as *ϕ* changes.

**Figure 16.**
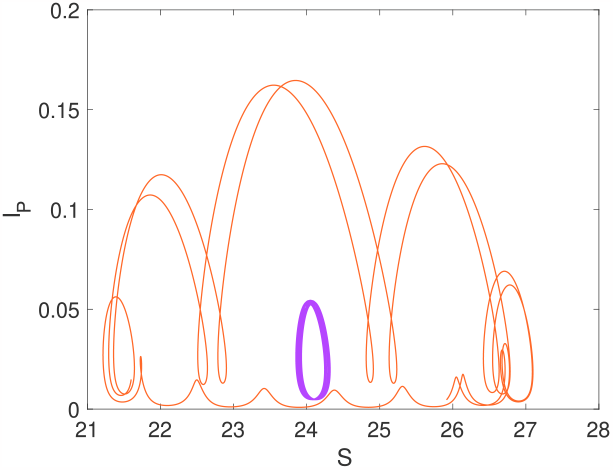
Phase space for seasonal model for two different values of *ϕ* = 1.46 (violet) and *ϕ* = 1.47 (orange) where *η* = 0.35, fixing other parameter from Table 2.

Figure 17 provides insight into the role of an unstable saddle limit cycle behaviour in the neighbourhood of the origin where *I*_*P*_ = 0 and *I*_*S*_ = 0 (see also the analysis of this phenomenon in [12]). Panels (a) and (b) of Figure 17 corresponds to *ϕ* = 1.43 and *ϕ* = 1.46, respectively, showing that the dynamics are bounded to a plane in the (*I*_*P*_, *I*_*S*_) space. Panels (c-d) depict the transition into chaotic behavior for *ϕ* = 1.47 and *ϕ* = 1.48674, respectively, illustrating the shape between one arbitrary return point of the projection of the torus dynamics in the (*I*_*P*_, *I*_*S*_) space.

**Figure 17.**
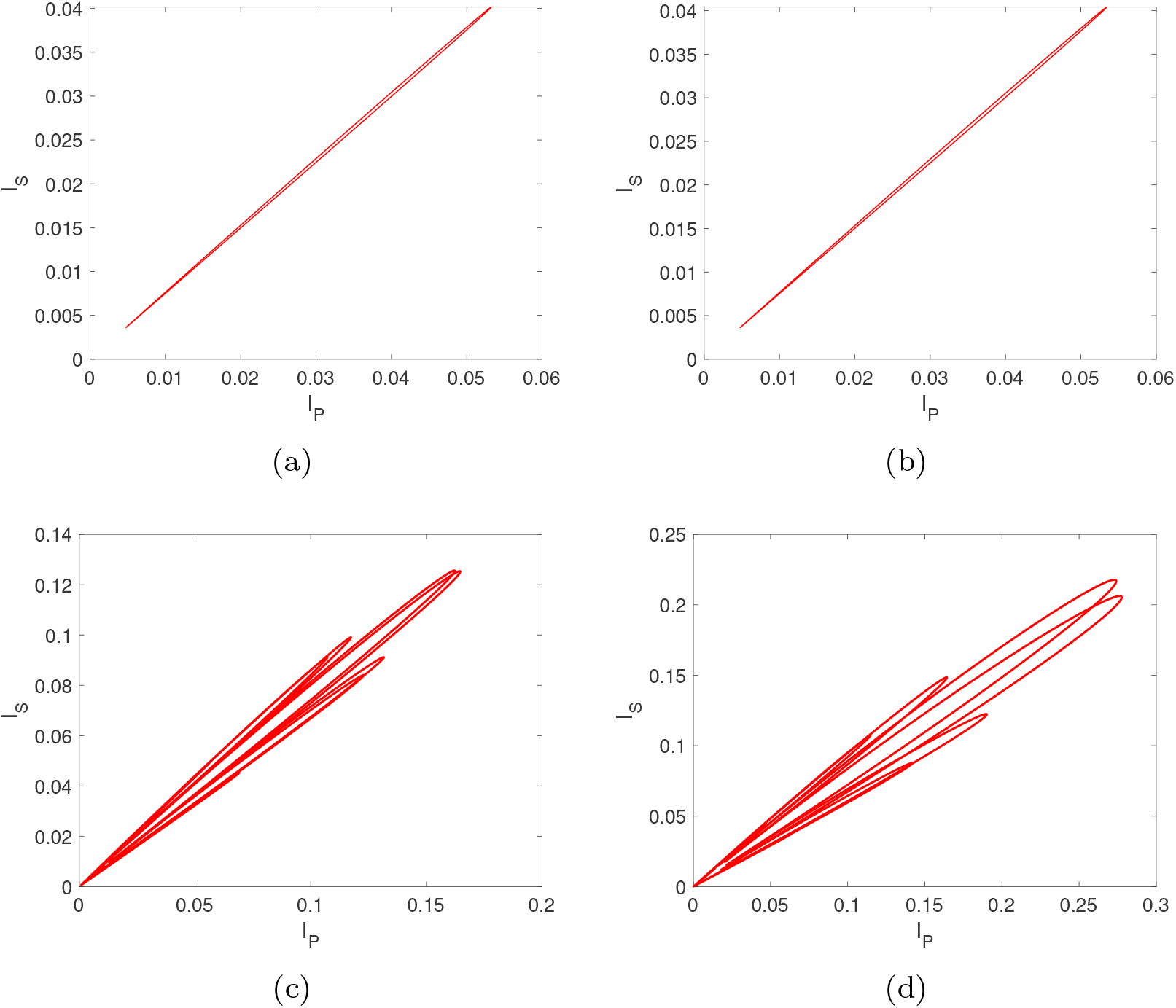
Projection of the trajectory on the *I*_*P*_, *I*_*S*_-plane for seasonal model for different *ϕ* values, and fixing other parameter from Table 2. (a) *ϕ* = 1.43, (b) *ϕ* = 1.46, (c)*ϕ* = 1.47, and (d) *ϕ* = 1.48674

These results indicate that the dynamics below the torus bifurcation point (*TR*) is characterized by a stable limit cycle, yet its shape exhibits erratic behavior.

## 6. Discussion and conclusions

The study of epidemiological scenarios characterized by chaotic dynamics represents a novel and crucial area of research, offering valuable insights into the dynamics of infectious diseases and contributing to the development of more accurate predictions for disease control strategies. Motivated by the epidemiology of dengue fever, a minimalistic two-strain mathematical model has revealed deterministic chaos in broader biological parameter regions than anticipated by previous models [5, 3, 6]. This model, incorporating a temporary cross-immunity period after primary infection with one serotype and disease enhancement in subsequent infections with different serotypes, has demonstrated complex dynamical behavior of the system.

More recently, a simplified version of the multi-strain dengue model proposed by Aguiar and colleagues [5] was studied in [34]. The authors investigated a compartmental model structured as a two-infection SIRSIR model without strain specificity of the pathogens. The model includes temporary immunity gained after a primary infection and incorporates an enhancement factor for a secondary infection based on the Antibody-dependent Enhancement (ADE) process, identifying rich dynamics and different bifurcation structures in the system. However, the explicit modeling of vector dynamics was not considered. In this study, we extend the SIRSIR model proposed in [34] by including explicit disease vector dynamics, leading to our SIRSIR-UV model. This extension provides insights into how disease vectors contribute to the overall system dynamics.

The stability analysis of the SIRSIR-UV model, using non-linear dynamical system theory, and the detailed bifurcation analysis conducted in this study are crucial steps in comprehending the qualitative behavior of the system. The identification of rich dynamical structures, including Bogdanov-Takens, cusp, and Bautin bifurcations, offers a deep understanding of the complexity inherent in the system’s behavior.

The recognition of a backward bifurcation for higher disease enhancement in secondary infections, exhibiting bi-stability of disease-free and endemic equilibria with a more extended biological temporary immunity period, is particularly important. This confirms that the combination of temporary immunity and disease enhancement significantly influences the complexity of the system dynamics, as first suggested in [7]. The formalization of the backward bifurcation using center manifold theory establishes a rigorous mathematical framework for analyzing this behavior. The computation of Hopf and global homoclinic bifurcation curves, along with the derivation of analytical expressions for transcritical and tangent bifurcations, enhances the depth of the epidemiological scenario analysis.

The observation of chaotic behavior after incorporating seasonal forcing highlights the importance of considering external factors, such as climate and weather patterns, that can influence disease spread. In the case of dengue fever, real data resembling chaotic behavior highlights the significance of including external forces resembling changes in mosquito abundance during the summer.

It is crucial to acknowledge that the absence of strain structure of pathogens may limit the model’s predictive power in certain scenarios. For instance, if multiple strains of a pathogen coexist, they may interact in complex ways not captured by a model assuming a single strain. Nevertheless, the SIRSIR-UV model provides a valuable starting point for exploring the dynamics of vector-borne diseases and identifying key factors driving their spread.

Our study has shown that the bifurcation structures identified in the SIRSIR-UV model closely align with and are comparable to those observed in the previously minimalistic SIRSIR model [34]. Consequently, this research offers valuable insights into the mathematical modeling of vector-borne diseases, underscoring the significance of simplifying assumptions, such as incorporating only implicit vector dynamics, particularly when constructing models without the application of vector control. These assumptions contribute to the manageability of mathematical analysis and modeling processes for complex models.

## Data Availability

There is no data used in this manuscript

https://www.example.com

## Acknowledgment

Akhil Kumar Srivastav acknowledge the financial support by the Ministerio de Ciencia e Innovación (MICINN) of the Spanish Government through the Juan de la Cierva grant FJC2021-046826-I. Maíra Aguiar acknowledges the financial support by the Ministerio de Ciencia e Innovacion (MICINN) of the Spanish Government through the Ramon y Cajal grant RYC2021-031380-I. This research is supported by the Basque Government through the “Mathematical Modeling Applied to Health” Project, BERC 2022-2025 program and by the Spanish Ministry of Sciences, Innovation and Universities: BCAM Severo Ochoa accreditation CEX2021-001142-S / MICIN / AEI / 10.13039/501100011033.

## Appendix

### 6.1. Calculation of ℛ_0_

To compute ℛ_0_, readers are directed to [37, section 4.5]. However, the specific calculations for this vector-host model are detailed below.

The basic reproduction number, ℛ_0_, for the system (1), is determined using the next generation matrix method outlined in [37]. The compartmental model is decomposed into two transition matrices, ℱ and, 𝒱 representing the inflow and outflow of the system, as follows:

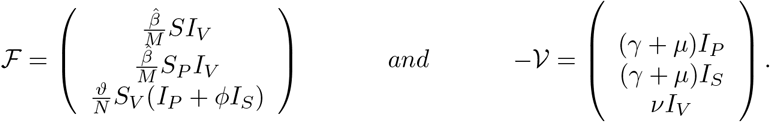

Given *F* as the matrix of partial derivatives of the transition matrix ℱ at the disease-free equilibrium *E*_0_,

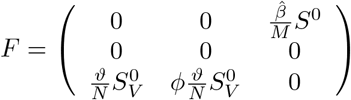

and *V* as the matrix of partial derivatives of the transition matrix 𝒱 at *E*_0_,

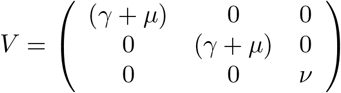

we calculate ℛ_0_ by constructing the next generation matrix *K* = *FV* ^−1^. Then, by solving the characteristic equation |*λ*− *K*|, we end up with a quadratic equation in terms of *λ*:

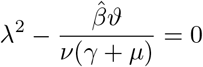

The quadratic equation has one non-negative eigenvalue. Therefore, ℛ_0_ is defined as the largest eigenvalue, in modulus, of the matrix *K* [37]. Thus,

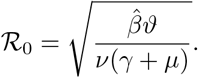

### 6.2. Proof sketch of Theorem 3.5

The proof follows the classical approach for establishing the stability of an equilibrium. The Jacobian matrix of the system (1) at the unique endemic equilibrium *E*_1_ is given by

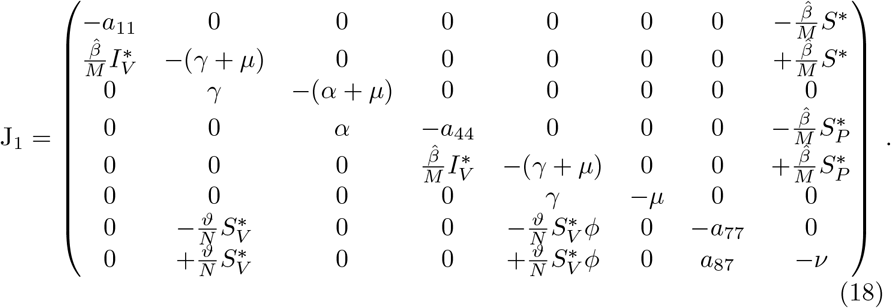

where

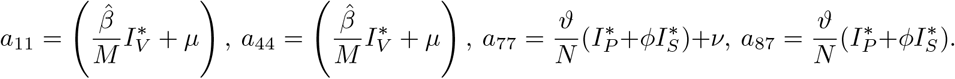

Moreover, the characteristic polynomial is determined, resulting in a polynomial of the same degree as the number of equations, which is 8 in this case. However, by decoupling the equations for the variables *S*_*V*_ and *R*, the size of the Jacobian is reduced by 2 dimensions. As a result, the characteristic polynomial has a degree of 6. Seeking solutions on the left side of the complex plane, one of the solutions is negative. The remaining solutions depend on three parameters and can be determined using Descartes’ rule or the Routh–Hurwitz criteria.

